# Evaluating large language models for predicting psychiatric acute readmissions from clinical notes of population-based EHR

**DOI:** 10.1101/2025.03.07.25323558

**Authors:** Terne Sasha Thorn Jakobsen, Enric Cristòbal Cóppulo, Simon Rasmussen, Michael Eriksen Benros

## Abstract

Psychiatric patients often have complex symptoms and anamneses recorded as unstructured clinical notes. Large language models (LLM) now enable large-scale utilization of text data; however, there is a current lack of LLMs specialized for psychiatric clinical data, as well as non-English data, haltering the application of LLMs across diverse clinical domains and countries. We present PsyRoBERTa: the first LLM specialized for clinical psychiatry, using population-based data with the currently largest collection of clinical notes of psychiatric relevancy (∼44 million notes) covering the eastern half of Denmark. The model was evaluated against three publicly available models, pretrained on either public general- or medical-domain text, and a baseline logistic regression classifier. Through extensive evaluations, we investigated the effect of domain-specific pretraining on predicting acute readmissions in psychiatric hospitals, explored important features, and reflected on (dis)advantages of LLMs. PsyRoBERTa succeeded in outperforming prior models (AUC=0.74), capturing information aligning with clinical practice, and additionally recognizing psychiatric diagnoses (AUC=0.85). This demonstrates the importance of domain-pretraining and the potential of LLMs to leverage psychiatric clinical notes for enhancing prediction of psychiatric outcomes.

## INTRODUCTION

Large language models (LLM) now enable large-scale utilization of the complex symptom descriptions and anamneses present within clinical notes from Electronic Health Records (EHR). However, at a large scale, clinical notes remain largely unutilized for psychiatric research and outcome prediction, with most efforts utilizing structured data from clinical studies, registries and EHR, as well as genetic data.^1–3^ This is partly explained by the fact that applying LLMs to clinical psychiatry data can be particularly challenging: Clinical text differs considerably, in vocabulary and structure, from the internet-originating text most general-purpose LLMs are based on, particularly regarding the many clinical abbreviations common in clinical EHR text. Moreover, LLMs are essentially large neural networks that have been trained to predict upcoming or missing words in a sequence of words, usually based on large corpora of internet-originating text. To apply an LLM for other tasks, such as predicting psychiatric outcomes from clinical notes, the LLM should also be *finetuned* specifically for this, which entails adding new learnable weights for the task, and backpropagating through the entire model to update the learned word representations (embeddings) according to the new purpose. Although a main advantage of LLMs is that they can be applied in such a *transfer learning* fashion, meaning they only need relatively brief finetuning for new tasks, because they have already learned good, general representations of words, there is substantial evidence suggesting LLMs should be further pretrained for the target domain (i.e. clinical psychiatry), before finetuning, to achieve best performance and parameter-efficiency.^4–9^

Several LLMs have been pretrained for the clinical domain with English EHR notes.^10–20^ A recent review^21^ identified 23 LLMs pretrained specifically on clinical text and found 17 of them to be trained on the same source data (MIMIC-III/IV),^22,23^ highlighting the challenge of attaining high-quality clinical text datasets and a current lack of diversity among available models. Furthermore, data in English is by far dominating the rapid development of clinical LLMs while development for other languages is slowly progressing.^4,5,24,25^ **Figure 1** gives an extended overview of LLMs pretrained on clinical notes^5,6,10–20,26–36^ and commonly evaluated against their base model, e.g. BERT,^37^ and models pretrained on biomedical research literature, e.g. BioBERT.^38^ The availability of LLMs for investigating mental health issues is currently much more limited,^39^ with most efforts focused on exploring mental health conditions in online fora using models such as MentalBERT^40^ trained on text from social media. So far, utilizing clinical notes for psychiatric outcome prediction has mostly built on other natural language processing (NLP) methods for information extraction, topic modelling, and sentiment classification for tasks such as predicting the use of mechanical restraint,^41^ psychosis symptom onset,^42,43^ and readmission.^44–47^ Recently, Perfalk et al (2024)^48^ evaluated machine learning (ML) models for predicting involuntary psychiatric admissions in the Central Denmark Region, such as a model trained with Bag-of-Words features (text represented by frequencies, disregarding order and context) from clinical notes. They highlighted the potential of a transformer-based model finetuned specifically for psychiatric clinical notes as a future direction more suitable for the sequential text data.

**Figure 1.**
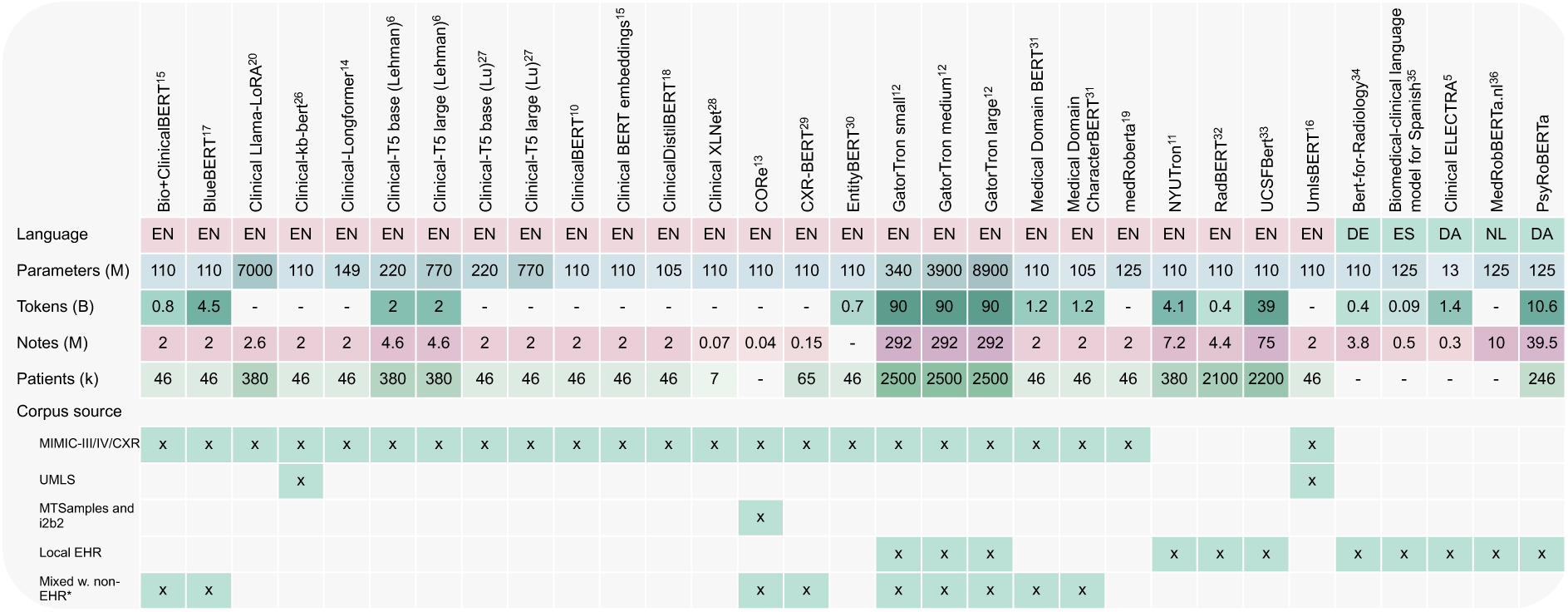
Overview of prior language models pretrained on clinical notes *Gradients of the colors signify volume, with darker colors for larger values within the property*.

Here, we present the first LLM specialized for clinical psychiatry using population-based data and the currently largest collection of clinical EHR notes with psychiatric relevancy. Our model, PsyRoBERTa, is pretrained on ∼40 million Danish clinical notes from 245,589 patients who have had contact with the mental health services. Clinical ML studies often only report a few performance metrics and no explainability or bias analyses. In this study, we conduct extensive evaluations when comparing our model to a logistic regression (LR) classifier (using a Bag-of-Words (BoW) representation of the frequency of unigram features) and three transformer-based LLMs pretrained on other sources and finetuned on our clinical notes. Pretraining LLMs is expensive and time-consuming, but it is a one-time expense that has the potential to enhance performance or reduce the amount of data needed for finetuning. With the task of predicting psychiatric acute readmissions as a case, we aim to answer the questions: 1) Does domain-specific pretraining enhance performance for predicting acute readmissions in psychiatry, as expected given previous research? 2) What features within clinical notes does an LLM find important for psychiatric acute readmissions and do these align with clinical practice? 3) What are the (dis)advantages of an LLM versus a simpler ML model?

## RESULTS

### An extensive and transparent evaluation framework

From Danish population-based electronic health records, we processed 43,996,617 clinical notes from 255,944 patients with at least one hospital contact with the mental health services between 2000 and 2022 (**Figure 2A**). Data preprocessing included: de-identification of patients and health professionals; splitting data based on randomized patient IDs to ensure independence of patients across train-validation-test-splits; de-duplication of content in clinical notes. A pretraining dataset and finetuning dataset was created, with patients within the finetuning test set excluded from the pretraining data (**Figure 2B**). The study then consisted of 1) pretraining PsyRoBERTa, 2) finetuning models, and 3) evaluation (**Figure 2C**). We pretrained PsyRoBERTa in four stages to observe the effect of continuous pretraining with more or lesser data. We then finetuned PsyRoBERTa and three publicly available Danish LLMs (MeDa-BERT,^49^ RøBÆRTa,^50^ and Danish BERT^51^, the latter hereon referred to simply as BERT) for predicting psychiatric acute readmissions and experimented with two datasets for finetuning, using either all notes within an admission (∼3.4M) or using only the discharge summaries (∼85k) to evaluate the difference between using the compressed information, i.e. discharge summaries, versus all notes. We reported performance of the LLMs and an LR baseline on eight metrics, with non-parametric bootstrapping confidence intervals, for a comprehensive representation of each model’s strengths and weaknesses. Due to class imbalance (23% cases), we emphasized Matthew’s Correlation Coefficient (MCC) as it is considered one of the most reliable metrics for imbalanced binary data.^52^ We further evaluated fairness across sex, age, region, and psychiatric diagnoses. Lastly, we explained learned patterns of PsyRoBERTa, by inspecting attention scores using the Attention Rollout method,^53^ and we explored the usefulness of its embeddings for psychiatric diagnosis recognition.

**Figure 2.**
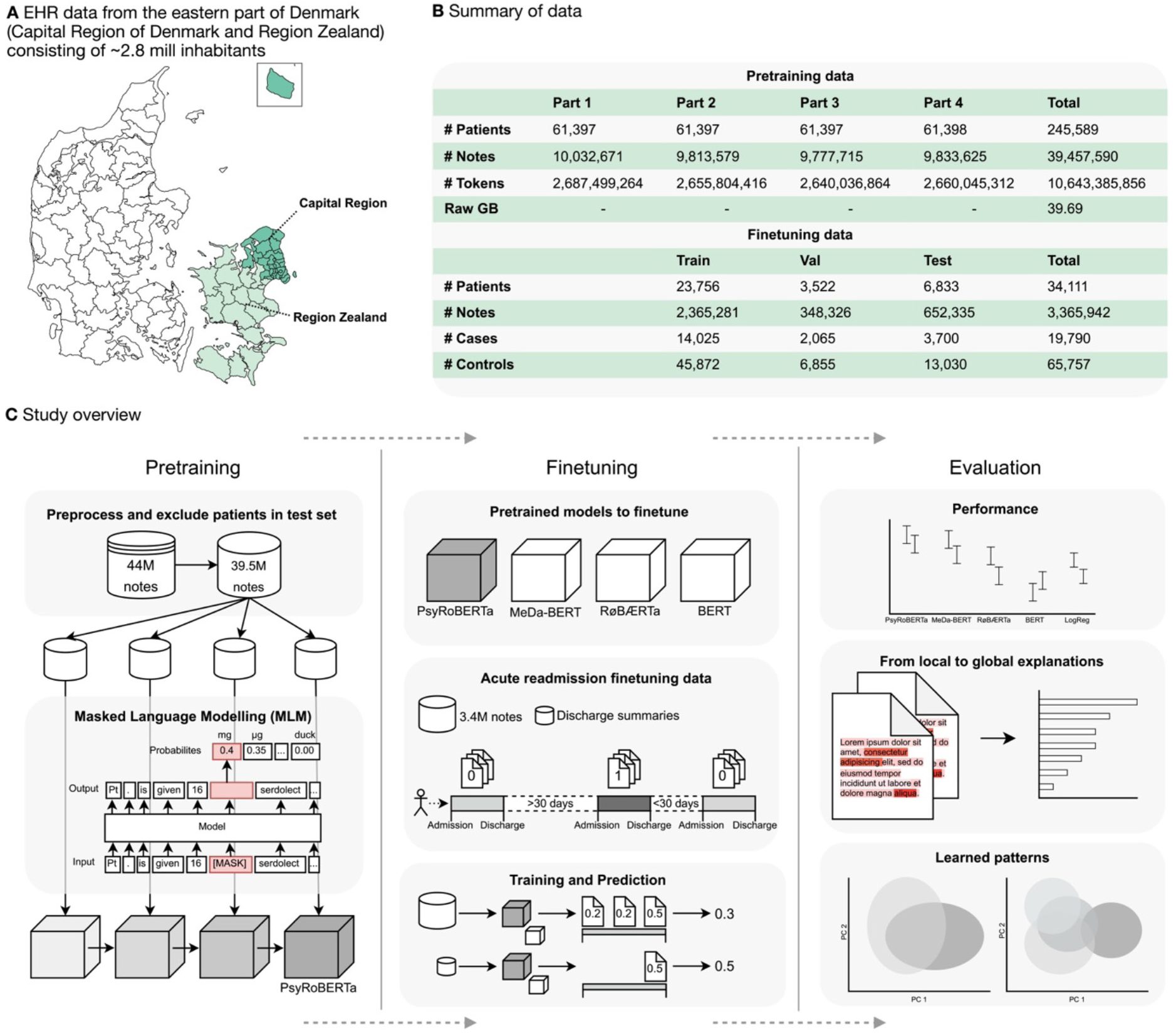
Graphical abstract with data description and study overview ***A.*** *Origin of EHR data. **B.** Size of pretraining and finetuning datasets. Raw GB refers to the size of a text file containing only the notes. Tokens were counted after tokenization with the RoBERTa tokenizer (byte-level BPE). Cases and controls refer to the admissions that lead or did not lead to acute readmissions. **C.** Study overview. This study consists of three steps: pretraining, finetuning, and evaluation. **Pretraining**: the clinical notes were preprocessed and patients within the test set of the finetuning data were excluded from pretraining. The remaining 39.5 million notes were split in four equally sized parts based on the number of patients with random selection. Starting with the RøBÆRTa model for MLM, we continued MLM training with each of the four data splits for 3 epochs, resulting in four models trained on incrementally more clinical notes. **Finetuning:** we finetuned four language models for predicting psychiatric acute readmissions; our model, PsyRoBERTa, and three publicly available models. Finetuning was carried out with two datasets, either containing all notes related to an admission or containing only the discharge summaries. An acute readmission was defined as an unplanned admission within 30 days after discharge. The models were trained to make a prediction for each textual input, with binary labels indicating whether the next admission was acute (1) or not acute (0)*, *and we took the mean over notes within each admission as the final predictions. **Evaluation:** We compared the predictive performance of each finetuned language model and a logistic regression classifier. We then retrieved explanations for PsyRoBERTa’s predictions and visualized embeddings to inspect learned patterns*.

### Domain-specific pretraining improved prediction of psychiatric acute readmissions

Our first question was whether domain-specific pretraining improves down-stream performance in our case as expected given the background literature. Overall, our model PsyRoBERTa consistently outperformed the non-clinical models on four metrics: Area Under the ROC curve (AUC), MCC, Area Under the Precision-Recall Curve (AUPRC), and weighted average F_1_ (**Figure 3C**). The second-best model was MeDa-BERT, which was pretrained on public clinical guidelines and medical websites,^47^ indicating that these data sources were also highly relevant. Comparing the performance of finetuning the models with discharge summaries versus all notes revealed that the latter gave the best performance and PsyRoBERTa was the best model in both scenarios. When finetuning with discharge summaries, PsyRoBERTa (MCC=0.285) had significantly better^48^ performance than all other models on the MCC metric: MeDa-BERT (MCC=0.264, p<0.01), RøBÆRTa (MCC=0.226, p<0.0001), BERT (MCC=0.215, p<0.0001) and LR (MCC=0.223, p<0.0001), and showed a 28% improvement from the LR baseline. When finetuning with all admission notes PsyRoBERTa (MCC=0.303) was still significantly better than RøBÆRTa (MCC=0.263, p<0.0001), BERT (MCC=0.193, p<0.0001) and LR (MCC=0.258, p<0.0001), and still better than MeDa-BERT (MCC=0.295), but by a smaller margin than previously, and showed a 17% improvement from the LR baseline. PsyRoBERTa, RøBÆRTa and MeDa-BERT were all excellent at correctly recognizing instances that did not lead to acute re-admissions (specificity: 91.9%, 94.8%, 93.1%, respectively) while recognizing positive cases was more challenging. However, PsyRoBERTa had a higher recall than the other language models, retrieving 33.2% of positive cases versus 30%, 23.7%, and 12.9% by MeDa-BERT, RøBÆRTa and BERT, respectively. We inspected the models’ predicted probabilities relative to the observed probabilities, i.e. true risk (also known as *calibration curves*, see **Figure S1**) and found that all LLMs tended to overestimate the risk of readmission but exhibited better calibration when finetuned on all notes. Comparing the performance of PsyRoBERTa and MeDa-BERT with the models they were continued pretrained from (RøBÆRTa and BERT) (**Figure 3B-C**) showed that the continued pretraining resulted in a 26% improvement in MCC of RøBÆRTa and a 23% improvement in MCC of BERT when finetuning with discharge summaries. When finetuning with all admission notes, we found a 15% improvement of RøBÆRTa and a 53% improvement of BERT (the improvement should be seen in the perspective that BERT performed worst overall). Furthermore, the difference in performance of finetuning with discharge summaries versus all notes indicated that there is additional relevant information beyond the summaries, but the improvement in performance was modest and potentially hampered by increased amount of irrelevant (noisy) information. One way to reduce noise in text data is to remove repetitions (duplication), and we found that our deduplication method, applied during preprocessing of the data, improved the performance of PsyRoBERTa by 2.4% while reducing training time by ∼3 hours.

**Figure 3.**
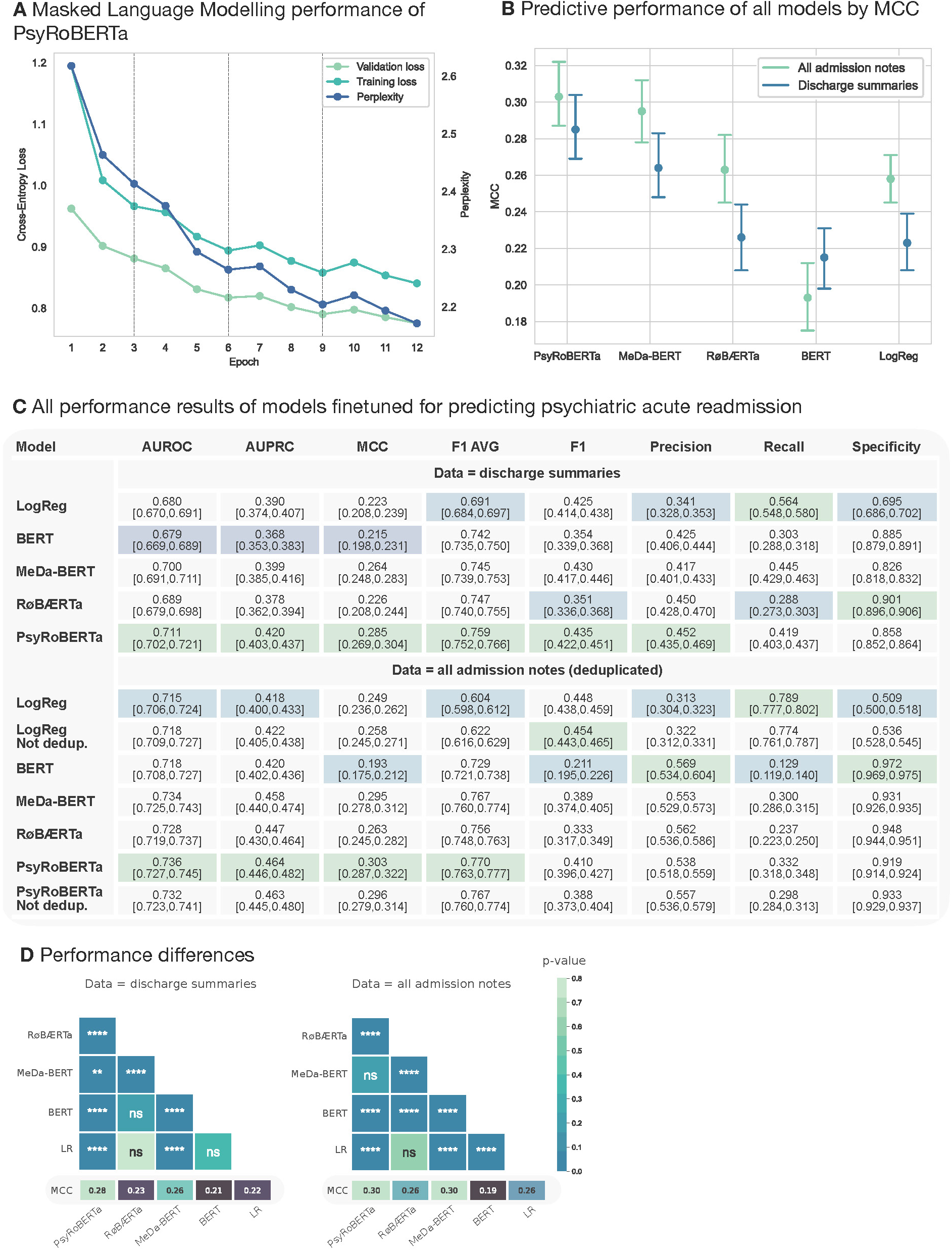
Model performance ***A.*** *Performance of the masked language modelling objective in pretraining. The striped line indicates the first epoch of the next model in our four-stage continuous pretraining. **B.** MCC of PsyRoBERTa, MeDa-BERT, RøBÆRTa, BERT and logistic regression. For each model on the x-axis, the left-most bar is the result from fine-tuning on all admission notes and the right-most is on discharge summaries. **C**. Performance of models fine-tuned for predicting psychiatric acute re-admissions. Top: results of finetuning with discharge summaries. Bottom: results of fine-tuning with all admission notes that are deduplicated, except the first and last row since it is not beneficial to deduplicate data for the logistic regression classifier (first row), and we show results of PsyRoBERTa when not deduplicating for comparison (last row). Green and blue highlights indicate best and worst score, respectively. 95% Confidence intervals are from non-parametric bootstrapping with 1000 resamples. **D.** Statistical significance for the observed difference in model performance by MCC. **p<0.01, ***p<0.001, ****p<0.0001*.

### Fairness analysis indicates performance consistency across diagnoses and patient demographics

PsyRoBERTa consistently outperformed all models on all demographic subgroups (**Figure S2**) as well as most diagnosis subgroups apart from ‘bipolar/manic’ and ‘other psychosis’ (**Figure S3**). Cross-group performance disparity was generally small for all models (**Figure S4**), but largest across the diagnosis subgroups since these had substantial differences in frequency and within-group class proportion (**Figure S5**). The difference in performance across diagnosis subgroups, measured as the difference between the largest and smallest AUC across groups, was smallest for PsyRoBERTa (0.111) and largest for MeDa-BERT (0.139), indicating greater fairness by performance parity of PsyRoBERTa. PsyRoBERTa performed best for patients with schizophrenia (AUC=0.750) or personality disorders (AUC=0.771) and worst for patients with anxiety (AUC=0.662) or depression (AUC=0.660) – the diagnoses with lowest proportions of acute readmissions. PsyRoBERTa (all admission notes) had good performance parity for all sex-, region- and age-groups (**Figure S6**), except for patients above 54 years of age (AUC=0.669). Similar results were observed for PsyRoBERTa finetuned on discharge summaries (**Figure S7-8**).

### Data size and training efficiency

We investigated performance improvements given increasing amount of pretraining data. We pretrained PsyRoBERTa on 39.5 million Danish clinical notes split in four parts to consecutively train the model for masked language modelling (MLM) for three epochs on each of the four parts of the pretraining data (i.e. three epochs with data part 1, then three epochs with data part 2 and so on, as visualized in **Figure 2C**). The MLM performance improved most in the first epochs, with an 8.4% improvement in validation loss from the first to the third epoch, and gradually diminishing gains throughout the pre-training process to 2.8% with the last part of the data (**Figure 3A**). During the validation of the four models on predicting acute readmissions from discharge summaries, the last model improved slightly faster (**Figure S9**). Consequently, using all available data for pretraining resulted in lowest validation losses but the gain of increasing training data was diminishing for each new part. After finetuning, we observed that the difference in predictive performance given discharge summaries versus all notes was smaller for PsyRoBERTa than the other models, with an absolute difference in MCC of 0.018 versus 0.031, 0.037, 0.022, and 0.035 for MeDa-BERT, RøBÆRTa, BERT and LR, respectively (**Figure 3B**). Furthermore, the difference in performance of PsyRoBERTa and MeDa-BERT was significant when training with discharge summaries but not when training with all admission notes. This could indicate that PsyRoBERTa needed less finetuning data for performance increases. To explore this further, we re-finetuned the three LLMs on 10-50% of the discharge summary dataset (**Figure S10**) and found that both PsyRoBERTa and MeDa-BERT had better onsets in predictive performance, reflecting more efficient learning of both domain-pretrained models. The difference between the onsets (i.e. whether PsyRoBERTa or MeDa-BERT had a better onset in predictive performance with less training data than the other), varied across the 10-50% of training data usage, and it was therefore not clear which, if any, was better in terms of needing less data for finetuning (see **Figure S10** for more details).

### Attention-based analysis identifies key clinical factors underlying model predictions

An advantage of LLMs over Bag-of-Words models is the ability to interpret important terms in each individual note (local patterns), which can provide fine-grained explanations of the prediction for individual patients. Due to the sensitive nature of the clinical notes, we did not analyze individual notes. Instead, we aggregated and sorted terms (sequences of words) by how much attention weight PsyRoBERTa gave them locally (using the method called Attention Rollout (AR)^53^) to show the most highly weighted bigrams (**Figure 4A**), trigrams (**Figure 4B**), and trigrams of instances where the model predicted acute readmission with high certainty (Softmax^54,55^ probability > 0.8). Through this, we identified five important categories of factors underlying model predictions: *psychosis*, *medication*, *level of function, alcohol and substances,* and *indications of a patient’s lack of insight into their own illness* (**Figure 4E**, see also supplementary Results, **Figure S11-13**). Comparing PsyRoBERTa’s AR scores to the coefficients of the LR classifier showed that both models recognized substance abuse as important (**Figure 4D**). However, LR (which was solely trained on unigram frequency and with no concept of word-order or time) seemed to rely most on terms describing a patient’s history of previous readmissions, as well as indications of violence and somatic medical issues.

**Figure 4.**
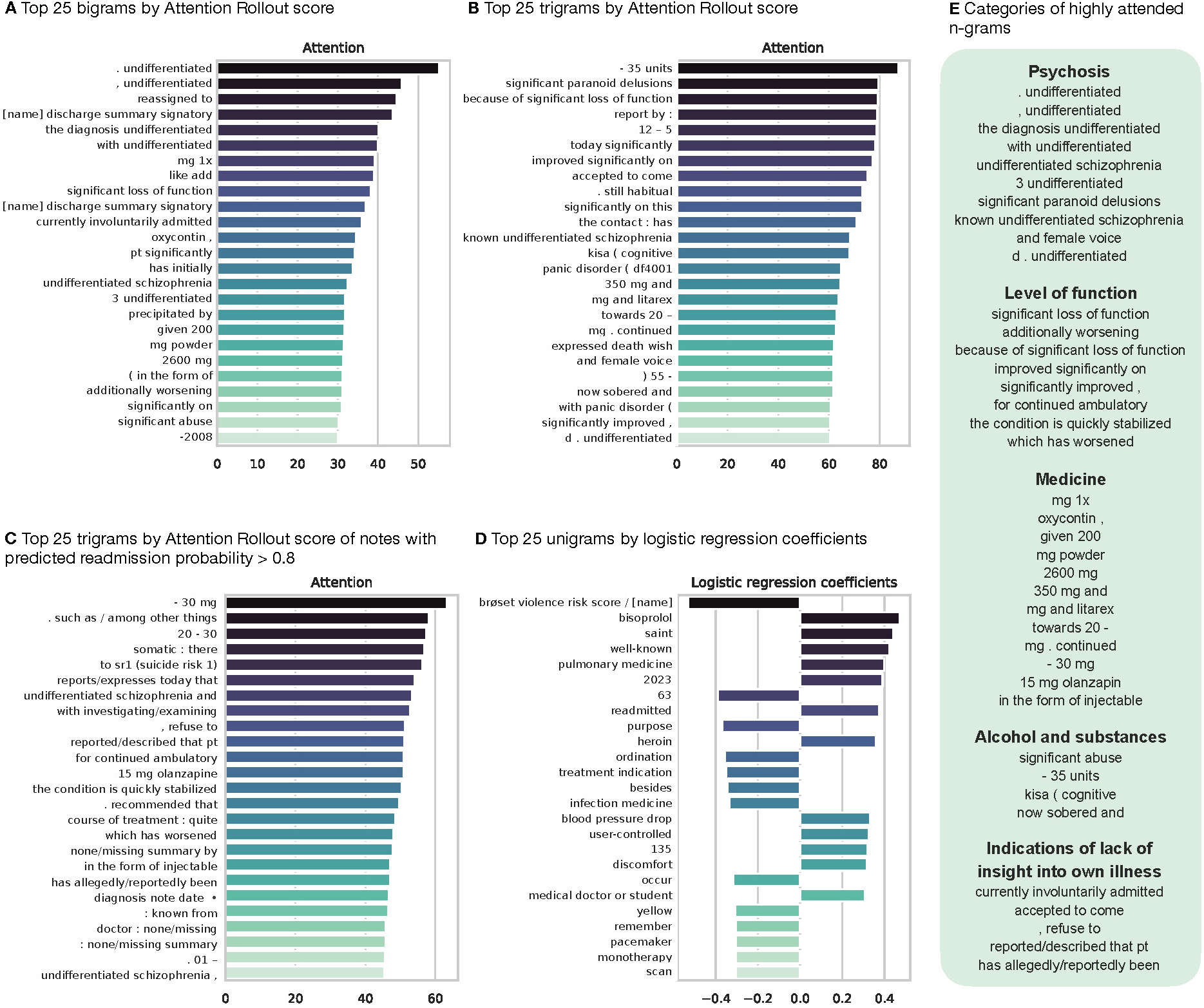
**Most highly weighted terms for psychiatric acute readmission prediction** *Top 25 most highly weighted bigrams (**A**) and trigrams (**B**) sorted by their max AR scores. N-grams were translated from Danish (**Figure S6-8**). Dashes indicate ambiguity and other possible translations. **C.** Top 25 trigrams of instances where the model predicted acute re-admission with high certainty, defined as a Softmax probability > 0.8. **D.** Top 25 terms by logistic regression coefficients. **E**. The highly weighted terms were categorized as related to psychosis, medicine, level of function, alcohol and substances, and indications of lack of insight into own illness. “undifferentiated schizophrenia” appeared in various forms among the most highly weighted n-grams. The word “significant” was prominent in descriptions of the patient’s level of function as well as descriptions of worsening and stabilization of the patient’s condition. Additionally, “significant” was highly weighted when describing “significant abuse” (likely substance abuse), and other descriptions of alcohol and substances included the number of units (of alcohol), being sobered and “kisa” (a treatment center specialized in stabilizing patients with alcohol and substance abuse). In the last category, “accepted to come” could indicate that the patient might not want treatment; the term “refuse” could be related to some patients refusing a diagnosis or treatment; the trigram “described that pt” could indicate that the patient (abbreviated as “pt”) was not themselves aware or able to describe an incidence; the term “allegedly” could indicate doubt in the veracity of the patient’s statement. The mean of embedded notes within an admission illustrated with PCA. **A**. The label indicates whether the next admission was acute (1) or not acute (0). **B**. The predicted probability of the next admission being acute. **C.** F0-9 are the 10 major psychiatric diagnosis codes. While patients may have more than one diagnosis, we show the current most severe diagnosis at the time of the admission. **D**. Most frequent subdiagnoses*.

### PsyRoBERTa’s representations of clinical notes enables feature clustering and diagnosis recognition

A beneficial side-effect of the contextual text representations, resulting from PsyRoBERTa, is that they can be used for other purposes than making predictions. With Principal Component Analysis (PCA), we explored PsyRoBERTa’s learned patterns and ability to recognize important aspects within the notes. We found many admissions resulting in acute readmissions located closely in the embedding space (**Figure 5A**) (however with substantial overlap of clusters) and the readmission risk spectrum learned by PsyRoBERTa was clearly visible (**Figure 5B, Figure S14-15**). Exploring other aspects with PCA revealed that the model captured the age spectrum (**Figure S16B**) and to some extent sex (**Figure S16A**) and note length (**Figure S17**). Most notably, PCA analyses revealed that PsyRoBERTa effectively separated admissions related to the 11 major psychiatric diagnoses (**Figure 5C**), indicating the ability to perform diagnosis recognition as a by-product of training the model to predict acute readmissions. Patients’ current diagnosis could thus be accurately recognized with a kNN classifier on pooled embeddings (AUC=0.832, MCC=0.489, k=50) (**Figure S18A**). The 7 most frequent sub-diagnoses were likewise well clustered (**Figure 5D**) and easily recognized (AUC=0.847, MCC=0.492, k=50) (**Figure S18B**).

**Figure 5.**
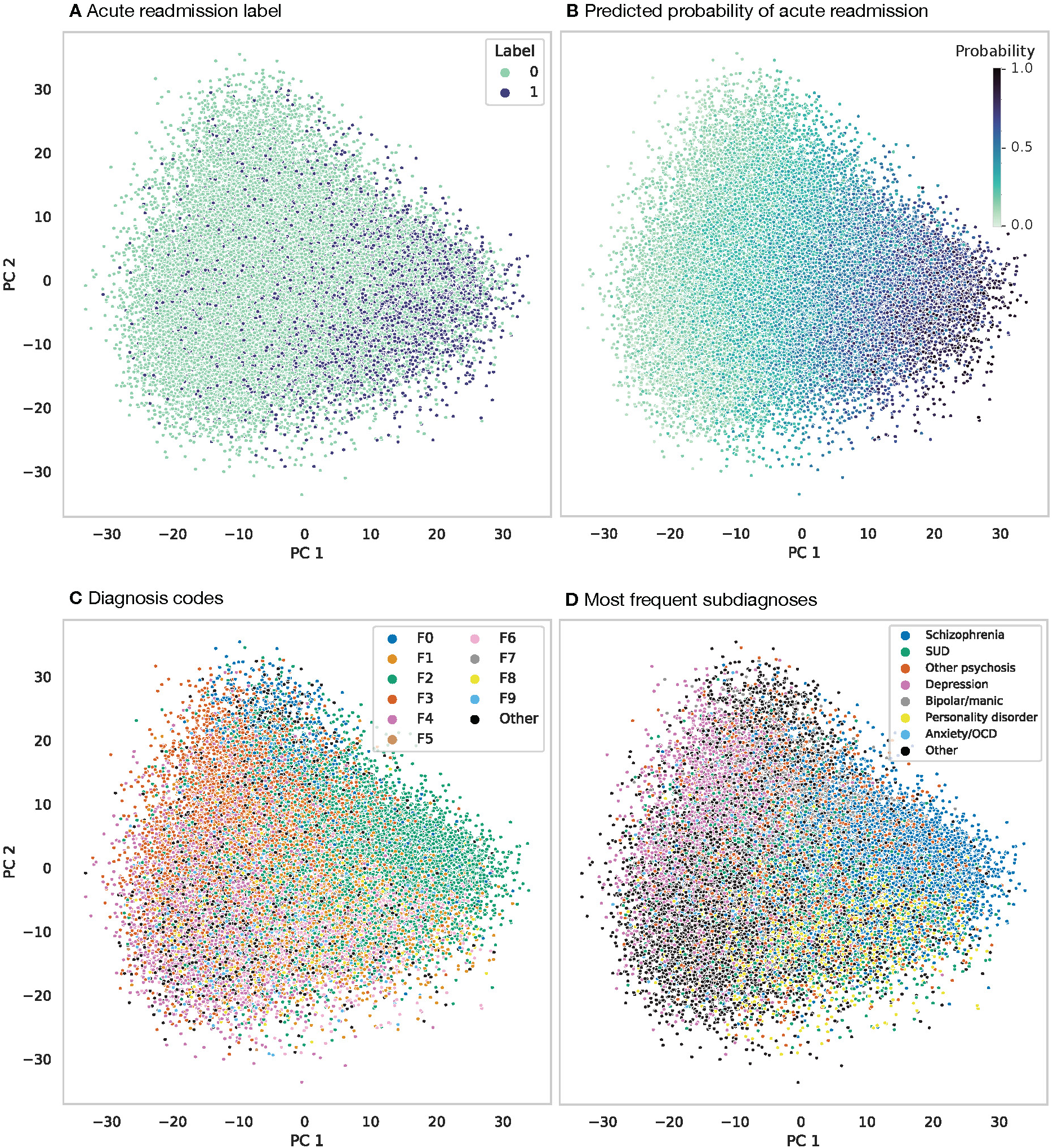
**Principal Component Analysis with mean pooled embeddings (hidden states) of the discharge summary finetuning trainset** *The mean of embedded notes within an admission illustrated with PCA. **A**. The label indicates whether the next admission was acute (1) or not acute (0). **B**. The predicted probability of the next admission being acute. **C.** F0-9 are the 10 major psychiatric diagnosis codes. While patients may have more than one diagnosis, we show the current most severe diagnosis at the time of the admission. **D**. Most frequent subdiagnoses.*

## DISCUSSION

We examined the effect of domain-specific pretraining by comparing our specialized model, pretrained on 39.5 million Danish clinical notes from 245,589 patients who have had contact with the mental health services, to those pretrained on more distant public domains, and we further analyzed data size effects in pretraining and finetuning. Our model, PsyRoBERTa, outperformed three public-domain LLMs and a logistic regression classifier on predicting psychiatric acute readmissions, with (AUC=0.736, MCC=0.303). Domain-specific pretraining with clinical notes enhanced performance by 15-26% in MCC. As expected, more pretraining, with more data, improved performance, and we found the gain of introducing new data to be diminishing for each new introduction, which is in line with previous research on pretraining RoBERTa-based models.^56,57^ We made versions of PsyRoBERTa pretrained on less data, to be used for tasks with rare outcomes. In line with the related work on domain-specific pretraining, our results indicate that the closer the pretraining data is to the target domain, the more the performance can be expected to improve, since the medically-informed model MeDa-BERT performed competitively to PsyRoBERTa, particularly when using a large finetuning dataset. Since RoBERTa is an upgrade from BERT, comparing the two models does not allow for making direct comparison of the impact of their respective pretraining data, and to do so, it would be necessary to train a PsyBERT and/or a MeDa-RoBERTa, which goes beyond our aim of comparing PsyRoBERTa to the relevant publicly available alternatives. Both PsyRoBERTa and MeDa-BERT exhibited more efficient learning during finetuning than the non-medical LLMs, meaning they require less data for finetuning for equal performance to non-medical models, which supports the need for domain-pretrained models for future clinical applications where data for finetuning is limited.^4,6,8,35,49,58,59^

PsyRoBERTa captured information about diagnoses and attended mostly to words related to psychosis, medication, the patient’s level of function, alcohol and substance abuse, and indications of lack of insight into own illness. Some of these factors, such as the patient’s level of function and insight into own illness, are not recorded in structured EHR and can therefore similarly not be captured by models built on structured data. Yet, these factors are closely related to clinical practice and gives insight to the clinicians’ evaluations of patients from direct interactions with them. With PCA and kNN we found that PsyRoBERTa embedded information about patients’ diagnoses and could therefore effectively recognize a patient’s current diagnosis in unseen notes, which can be useful for making clinicians aware of diagnoses missed or unrecorded in the structured EHR data.

The simple LR classifier also recognized substance abuse as an important factor but gave more weight to a history of prior (re)admissions. we speculate that while prior readmissions are an important predictive feature easily captured by the LR classifier, more complex factors are better captured by LLMs. An important distinction between LR and PsyRoBERTa, that could explain this, is that learned coefficients of LR are static while PsyRoBERTa’s weighing of terms is dynamic or *contextual*.^37^ This means that the occurrence of ‘bisoprolol’ (medicine for high blood pressure), with a positive LR coefficient, will always count towards an increased risk of readmission by LR. PsyRoBERTa instead takes the context into account and gives different weight to the word accordingly. This is a great advantage since such a feature is likely to be spurious if given equal weigh regardless of context. However, context-dependence makes interpretability of general associations between features and predictions challenging, and learning good contextual representations requires substantially more model training. If a modest performance difference does not justify the increased complexity and cost of LLMs for a specific project, there are other advantages that might: We have shown that the contextual, high-dimensional representations also allow for other ways of investigating patterns learned by the model and can be multi-purpose without additional training.

Our results indicated that predicting hospital readmissions is challenging, and related studies with English text data have reported similarly modest performance with AUC scores of 0.714,^10^ 0.726,^60^ and 0.798^11^ for predicting ICU readmissions, psychiatric readmissions, and all-cause readmission, respectively. On the related task of predicting psychiatric *involuntary* admissions (which is expected to be somewhat easier as it involves a specific subgroup of patients and is likely less influenced by early discharges due to resource scarcity), Perfalk et al. (2024) reported AUCs of 0.76 with an ML model (XGBoost).^48^ They proposed finetuning language models as a future direction to enhance performance compared to ML models. We found that finetuning general-domain LLMs (RøBÆRTa and BERT) did not result in significant increase of performance compared to a simple LR classifier, and hence it is crucial to apply an LLM pretrained with clinical data.

Limitations of this study includes the difficulty of providing robust explanations for LLMs’ predictions.^61–63^ The method used in this study, Attention Rollout, peeks into the attention mechanism, which is fundamental for transformer-based LLMs, however, it only partly explains the model’s decision-making.^64^ Furthermore, the LLMs used in this study were architecturally constrained to an input length that necessitated splitting some notes into smaller chunks and methods for encoding longer sequences could be a promising future direction.^14^ We did not experiment with large generative language models (such as GPT, Llama, Gemini) because of their limitations: Closed-source generative models are currently not permissible for use with patient data due to privacy concerns, and open-source generative models are computationally expensive, their predictions are sensitive to prompt formulations, and recent studies have found that smaller task-specific RoBERTa-based models perform better at classification than large generative models.^65–67^

We have presented the first large language model specialized for psychiatric outcome prediction using clinical notes, with psychiatric acute readmissions as proof-of-concept. The model outperformed publicly available alternatives and achieved state-of-the-art performance in predicting readmission from Danish clinical notes, with comparable performance to related work in English. We showed that our model relied on factors related to every-day clinical practice and which are not found in structured EHR data. Furthermore, the model excelled at diagnosis recognition without further training, highlighting the multi-purpose nature of LLMs. Importantly, PsyRoBERTa can be finetuned for many clinical use cases with the expectation of better performance and accelerated learning compared to non-clinical Danish LLMs.

## METHODS

### Study population and electronic health record data

Our data consists of 43,996,617 clinical notes from Danish population-based electronic health records covering around half of the Danish population (the Capital Region of Denmark and Region Zealand). The clinical notes were from 255,944 patients with at least one hospital contact with the mental health services between 2000 and 2022. The data consists of all the patients’ clinical notes and was thus not limited to contact with psychiatry. We de-identified the notes by masking patients’ personal and identifiable information and, additionally, developed a method for masking health professionals’ names.

### De-identification

Patients’ personal and sensitive information, including patients’ names, relatives’ names, addresses, social security numbers, phone numbers, and email addresses were first masked by using Python regular expressions. Further masking of (unknown) names of doctors and other health professionals was performed with regular expressions and the use of lexicons derived from online resources.

### Pretraining data

To ensure that our pretrained model could be used for unbiased evaluations of future downstream tasks, we split the data, based on randomized patient IDs, into four parts with approximately equal number of (non-overlapping) patients. This enabled pretraining in four stages with an increasing amount of data. Notes were concatenated and split to the maximum sequence length of the model and patients in the validation and test sets for our task were removed from the pretraining data. This resulted in a total of 39.5 million notes for pretraining, with 5% of these patients for MLM validation.

### Finetuning data

Finetuning was based only on the clinical notes from psychiatric hospital admissions. Train, validation, and test set splits (70/10/20) were based on randomized patient IDs. Binary labels indicated whether the next admission was acute (1) or not acute (0), with acute defined as unplanned and within 30 days after discharge. There was high class imbalance, with 19,790 cases (23%) and 65,757 controls (77%). We finetuned with datasets consisting of either 1) only the *discharge summaries* (∼86 thousand), or 2) *all admission notes* (∼3.4 million). For the latter, we additionally applied a deduplication method (see below). Long notes were split in chunks of the LLMs maximum input length but when finetuning on all admission notes, we concatenated notes within an admission, adding a separator token to signal the start of a new note within the text, before splitting.

### De-duplication

De-duplication of notes followed the DedupCont method of Liu et al. (2022)^68^ except that we only deduplicated within encounters (admissions) rather than across all notes. For each encounter, all notes were split on line-breaks (\n) into chunks. A chunk was removed if it was an exact match to any prior chunks in the encounter, and the chunks remaining were concatenated.

### Pretraining of Psychiatric Clinical RoBERTa model (PsyRoBERTa)

We initialized PsyRoBERTa with weights from RøBÆRTa,^50^ a RoBERTa-base^56^ architecture consisting of 12 transformer encoder blocks and self-attention heads, a hidden size of 768, a vocabulary size of 50265 tokens, and a total of 125 million parameters. RøBÆRTa was pretrained on the Danish part of the multilingual MC4 dataset.^69^ With our clinical notes, we continued training towards the masked language modelling (MLM) objective – a standard method for pretraining – predicting tokens that were randomly masked with a 15% probability. This pretraining was carried out in four consecutive stages, training for three epochs on each of the four pretraining data parts and resuming from the checkpoint of the previous model. The entire pretraining took ∼62 hours and was carried out using HuggingFace’s Accelerate library and on a Microsoft Azure Virtual Machine with 4 NVIDIA A100 GPU’s (Standard_NC96ads_A100_v4) with a cost of $19.10 per hour. See all hyperparameters in **Table S1**.

### Finetuning language models

To enable outcome prediction, a language model pretrained with MLM must be finetuned (trained) for the specific task. We finetuned PsyRoBERTa and three publicly available Danish LLMs (Danish BERT,^51^ RøBÆRTa,^50^ and MeDa-BERT^49^), for predicting psychiatric acute readmission, using either discharge summaries or all admission notes. In this process, a linear classification head was added to each model and during training both the classification head and the LLM’s weights were optimized for the prediction task, resulting in models that output a probability of the next admission being acute. All models were finetuned for 12 epochs and with equal hyperparameters (**Table S1**). The number of epochs was chosen based on inspecting the recline in training and validation losses (**Figure S19**) and with consideration for that finetuning should be cost-effective. We had chosen 12 as the maximum although the losses were persistently reclining and may have improved with more epochs, and we used the same number for all models for fair comparison. Due to high class-imbalance, we rescaled the weight given to each class in the cross-entropy loss function. The finetuning was carried out using PyTorch and Huggingface on a Microsoft Azure Virtual Machine with 4 NVIDIA A100 GPU’s (Standard_NC96ads_A100_v4) with a cost of $19.10 per hour and it took ∼14 hours with all admission notes and ∼1.35 hours with discharge summaries for each of the four language models (BERT, MeDa-BERT, RøBÆRTa, PsyRoBERTa).

### Logistic regression classifiers

The text was lowercased and some punctuation removed before training LR classifiers on the 5000 most frequent unigrams, using L2 regularization and balanced class weights. We used grid search to find the best feature representation for the classifiers (optimizing average precision): we searched two vectorizers (count and TF-IDF, see below) and n-gram ranges of (1,1), (1,2), (1,3) and (1,4), where a range of (1,4), for instance, means including both unigrams, bigrams, trigrams, and 4-grams. The vectorizers represent text in a Bag-of-Words style (where the text is represented by frequencies, disregarding order and context). The count vectorizer represents each note as a vector with counts of token occurrences, and Term Frequency-Inverse Document Frequency (TF-IDF) additionally re-weighs each token by its frequency within all notes in training set. The best feature representation of discharge summaries was counts of unigrams and the best for all admission notes was TF-IDF of unigrams.

### Evaluating finetuned models

While there can be numerous clinical notes during a hospital admission, the LLMs provide predictions *per text* and some notes were split into several pieces of text. The LLMs outputs are Softmax^54,55^ probabilities (between 0 and 1) for each text. To provide a single predicted probability of acute readmission, we took the mean of the texts’ Softmax probabilities for the positive class for each admission. We evaluated predictive performance on a wide range of metrics and computed 95% confidence intervals with non-parametric bootstrapping using 1000 resamples. P-values were also calculated with bootstrapping (1000 resamples and 95% confidence level), calculating differences in the metric (MCC and AUC) of two models and computing the p-value for the null hypothesis that there is no difference in the performance. PsyRoBERTa was evaluated against three public LLMs to show the performance when pretraining on millions of clinical notes is not possible. However, the public models were also finetuned on clinical notes since they cannot predict outcomes out-of-the-box. We emphasized Matthews Correlation Coefficient (MCC) given its greater reliability for binary classification with imbalanced data.^52^ The deduplication method was only evaluated on PsyRoBERTa and the LR classifier.

### Explainability

We explained the predictions of PsyRoBERTa with Attention Rollout (AR),^53^ which attributes a score (attention weight) to each input token indicating the token’s importance for the prediction *locally* (i.e. specific for the input-prediction pair). Since weights were assigned to tokens of the RoBERTa tokenizer, which sometimes splits words into several tokens, we merged tokens and their weights to reconstruct the complete words. Additionally, we experimented with either using the mean or max of the weights when merging. Then, *to approximate global explanations*, we summed weights given to unique, lowercased words and normalized them by their frequency before inspecting highly weighted words. We focused the analysis on the 10% most frequent words, since infrequent words could be highly weighted but less relevant overall, and we only inspected words that were used in notes of at least three patients. We inspected highly weighted unigrams (1-word sequence), bigrams (2-word sequences) and trigrams (3-word sequences), including punctuations as “words”. Treating LR coefficients as global explanations for this model, we compared highly weighted terms of LR to the AR scores.

### Clustering and recognition

The transformation of clinical notes to multi-dimensional embeddings with finetuned PsyRoBERTa creates representations of words that are dynamic (meaning the representation changes depending on the context) and captures semantic knowledge (such that words used in similar contexts will have more similar representations). We used principal component analyses (PCA) to visualize these high-dimensional embeddings of PsyRoBERTa finetuned on discharge summaries. (Additional visualizations with UMAP in **Figure S20-23**). We performed inference on the train and test set of discharge summaries and mean pooled the last hidden state. We then further mean pooled the several splits of notes, that may exist within an admission, to finally have one vector representation of each admission. We reduced the vectors for the train set to two dimensions with PCA and visualized the embedding space in relation to seven variables: true class, predicted probability, predicted class, sex, age, amount of note splits, and main psychiatric diagnosis. k-nearest neighbors (kNN) classifiers were trained and tested on the vectors, with k={5,10,50}, to evaluate the model’s ability to cluster and recognize these important variables.

### Fairness and bias

The term *fairness* is inherently value-based, and there exists several definitions that can appear conflicting.^70,71^ In this study, we evaluate *group fairness*, which we define as non-discrimination for socio-demographic groups. This is rooted in the ideal that clinical prediction models should not discriminate based on attributes such as age, sex/gender, race/ethnicity and socio-economic status.^72^ We evaluated group fairness through the lens of *equalized odds* (equality in true positive and true negative rates) and *performance* parity.^73,74^ Furthermore, the explainability methods described above were used to reveal and better understand potential biases. (Un)fairness and bias are connected phenomena, since unintentional biases can result in disparate performance across groups. We investigated the demographic attributes sex (binary), four age groups (below 18, 18-34, 35-54, and above 54 years of age) and region (Capital Region and Region Zealand). Ethnicity, although a relevant factor for a bias analysis, was unfortunately underreported. We further investigated fairness across the most frequent (sub)diagnoses: schizophrenia, other psychosis, substance use disorder (SUD), depression, bipolar/manic, personality disorder, and anxiety/obsessive compulsive disorder (OCD).

### Data and ethics approvals

This project is part of the PRECISE project where the overall aims is to utilizing health data to increase the understanding of mental disorders and to pave the way for improvements of current treatments for mental disorders. The project is approved by the Data Protection Agency of the Capital Region of Denmark and the Danish Society for Patient Safety (approval numbers: P-2020-101 and R-22002033). According to Danish regulations, the analyses do not require informed consent. All data are available to researchers in the regions after the relevant approvals from the Danish Health authorities. All patient data were pseudonymized and, as far as possible, only analyzed as summary data consisting of at least 3 patients in each group.

## DATA AVAILABILITY

The data used in this study are not publicly available to ensure the security and compliance with patient privacy regulations. Data can only be available for those researchers that meet the criteria for access to the confidential data, to be used in the work for improving the evidence base and quality in the patient treatment. The data and model cannot be shared openly according to Danish regulations due to potential personal identifiable information but is accessible for researchers with appropriate legal permissions on secured servers.

## CODE AVAILABILITY

The non personally identifiable part of the code will be shared on www.github.com/terne/PsyRoBERTa.

## ACKNOWLEDGEMENTS

The work was supported by unrestricted grants from the Lundbeck Foundation (grant number R278-2018-1411 and R383-2022-285) and from the Mental Health Services of the Capital Region of Denmark. S.R. was supported by the Novo Nordisk Foundation (NNF23SA0084103).

## AUTHOR CONTRIBUTIONS

Dr Benros had full access to all of the data in the study and takes responsibility for the integrity of the data and the accuracy of the data analysis. Drs Rasmussen and Benros are considered co–senior authors.

*Concept and design:* Jakobsen, Rasmussen, Benros.

*Acquisition, analysis or interpretation of data:* Jakobsen, Cóppulo, Rasmussen, Benros.

*Drafting of the manuscript:* Jakobsen, Benros.

*Critical review of the manuscript for important intellectual content:* All authors.

*Statistical analysis:* Jakobsen.

*Obtained funding:* Benros, Rasmussen,

*Administrative, technical, or material support:* Cóppulo, Benros.

*Supervision:* Rasmussen, Benros.

