## Supplementary for "Evaluating large language models for predicting psychiatric acute readmissions from clinical notes of population-based EHR"

### SUPPLEMENTARY INFORMATION

This file includes:

Results:
- Explaining predictions
- Embedding LLM results

Tables:
- Table S1. Hyperparameters for pretraining and finetuning.

Figures:
- Figure S1. Calibration curves
- Figure S2. Demographic fairness
- Figure S3. Diagnosis fairness
- Figure S4. Min-max difference in performance across subgroups
- Figure S5. Diagnosis fairness analysis of PsyRoBERTa finetuned on all admission notes
- Figure S6. Demographics fairness analysis of PsyRoBERTa finetuned on all admission notes
- Figure S7. Diagnosis fairness analysis of PsyRoBERTa finetuned on discharge summaries
- Figure S8. Demographics fairness analysis of PsyRoBERTa finetuned on discharge summaries
- Figure S9. Impact of increasing pretraining data on the downstream task of predicting acute readmissions from discharge summaries
- Figure S10. Language model data requirement analysis
- Figure S11. Top 25 most highly weighted bigrams and trigrams by Attention Rollout score
- Figure S12. Danish and English translations of the most highly weighted ngrams with their mean and max AR scores
- Figure S13. Danish and English translations of the most highly weighted ngrams with their mean and max AR scores (high risk instances)
- Figure S14. Visualising label and prediction clusters with PCA
- Figure S15. Acute readmission prediction performance of kNN classifiers
- Figure S16. Visualising sex and age clusters with PCA
- Figure S17. Visualising clusters of the number of splits of discharge summaries (from data preprocessing) with PCA
- Figure S18. Diagnosis clusters visualized with PCA and diagnosis recognition performance of kNN classifiers
- Figure S19. Training and validation loss of language models finetuned for predicting acute readmission
- Figure S20. Class clusters visualized with UMAP
- Figure S21. Prediction clusters visualized with UMAP
- Figure S22. Diagnosis clusters visualized with UMAP
- Figure S23. ROC and Precision-Recall curves of language models
- Figure S24. AUPRC and weighted $F_1$ of finetuned language models and logistic regression

#### Supplementary Results

**Explaining predictions**

PsyRoBERTa attention weights were analyzed with Attention Rollout (AR).[1] We sorted terms by their AR scores and explored the top 25 most highly weighted bigrams (**Figure 4A**) and trigrams (**Figure 4B)** and found that most of the highly weighted terms were related to four categories: *psychosis*, *medicine*, *level of function*, and *alcohol and substances*. "Undifferentiated schizophrenia" was a particularly prominent term, appearing in various forms among the most highly weighted bigrams and trigrams. Terms related to medicine included dosages such as "2600 mg", specific drugs such as "oxycontin", and indications of raising or continuing the dosage. The word "substantial" was often used of the patient's level of function, describing "substantial loss of function" and "substantial improvement", as well as descriptions of worsening and stabilization of the patient's condition. Additionally, "substantial" was highly weighted when describing "substantial abuse" (likely substance abuse), and other descriptions of alcohol and substances included the number of units (of alcohol), being sobered and "kisa". The latter being a reference to a treatment center specialized in stabilizing patients with alcohol and substance abuse.

When exploring the top 25 trigrams of instances where the model predicted acute re-admission with high certainty, which we defined as a SoftMax probability larger than 0.8, a fifth category emerged: indications of a patient's lack of insight into their own illness (**Figure 4C**). The trigram "accept to come" could indicate that the patient might not want treatment; the term "refuse" could be related to some patients refusing a diagnosis or treatment; the trigram "described that pt" could indicate that the patient (abbreviated as "pt") was not themselves aware or able to describe an incidence; the term "allegedly" could indicate doubt in the veracity of the patient's statement, such as when a patient experiencing psychosis describes having been somewhere or done something impossible.

Danish version, translations and corresponding attention weights are found in **Figure S11-13**.

**Embedding LLM results**

We experimented with using the embeddings of PsyRoBERTa for prediction *without finetuning the embeddings*. We froze the LLM layers and trained only the parameters of the linear classification head (on the discharge summary dataset) for 12 epochs (which was the same number of epochs used in our finetuning experiments). The performance of this "Embedding LLM" for predicting psychiatric acute readmissions was found to be near random (AUROC=0.596 [0.585,0.606], AUPRC=0.284 [0.272,0.297], MCC=0.044 [0.028,0.063], $F_1$ weighted average=0.692 [0.684,0.702], $F_1$=0.041 [0.032,0.05]), highliting the importance of finetuning embeddings for the given task.

#### Table S1. Hyperparameters for pretraining and finetuning

| Parameter | Pretraining | Finetuning |
|---|---|---|
| Batch size (per device) | 64 | 64 |
| Epochs | 3 (12 total) | 12 |
| Total steps | 243,612 | – |
| Seq. length | 512 | 512 |
| Optimizer | AdamW | AdamW |
| Weight decay | 0.0 | 0.01 |
| LR | 5e-5 | 3e-6 |
| LR decay | linear | On platau |
| Warm up steps | 1000 | 0 |
| MLM probability | 0.15 | – |
| Gradient accum. steps | 1 | 1 |
| Mixed precision | fp16 | fp16 |

Pretraining and finetuning was performed with mainly default hyperparameters. Batch size was chosen by the largest possible for our available resources. For finetuning, we used a low learning rate of 3e-6 after finding larger learning rates [1e-3, 3e-5, 1e-5] unsuccessful for predicting beyond the majority class on the validation set with PsyRoBERTa and RøBÆRTa.

#### Figure S1. Calibration curves

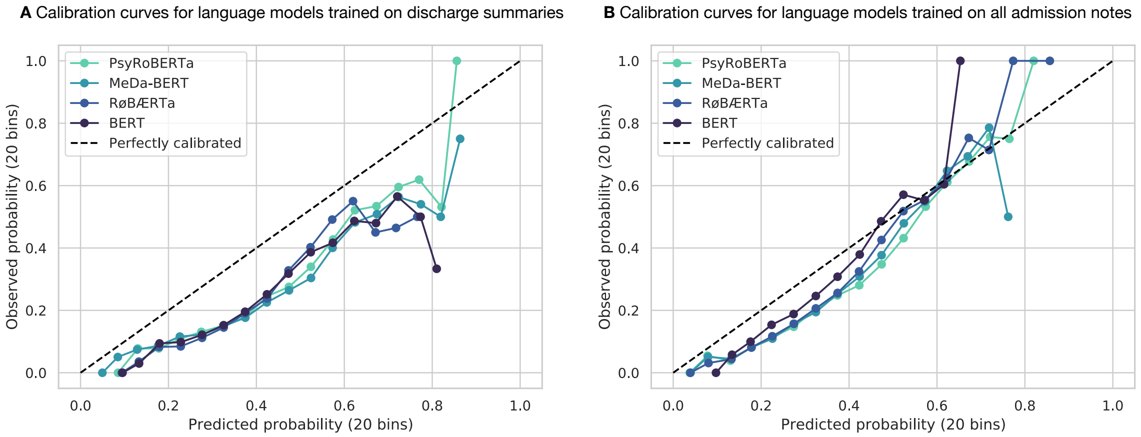


Calibration curves of language models finetuned on discharge summaries (**A**) and all admission notes (**B**). All LLMs tended to overestimate the risk of readmission but exhibited better calibration when finetuned on all notes. Finetuning with discharge summaries led to an overestimation of the probability of readmission throughout the whole threshold spectrum. However, finetuning with all admission notes overestimated the probability of readmission when the observed probability was below approximately 0.6 but tended to underestimate the probability above that threshold.

### Figure S2. Demographic fairness

| Model | TPR | TNR | AUROC | TPR | TNR | AUROC |
|---|---|---|---|---|---|---|
| | Data = discharge summaries | | | Data = all admission notes (deduplicated) | | |
| **Sex = Female** | | | | | | |
| LogReg | 0.718 [0.707, 0.729] | 0.519 [0.494, 0.543] | 0.665 [0.650, 0.680] | 0.564 [0.552, 0.575] | 0.743 [0.722, 0.764] | 0.711 [0.696, 0.724] |
| BERT | 0.907 [0.900, 0.914] | 0.266 [0.244, 0.290] | 0.676 [0.661, 0.691] | 0.973 [0.969, 0.977] | 0.106 [0.093, 0.120] | 0.708 [0.695, 0.722] |
| MeDa-BERT | 0.848 [0.839, 0.856] | 0.423 [0.400, 0.450] | 0.693 [0.677, 0.708] | 0.941 [0.935, 0.946] | 0.259 [0.238, 0.280] | 0.727 [0.713, 0.741] |
| RøBÆRTa | 0.911 [0.905, 0.918] | 0.253 [0.232, 0.276] | 0.678 [0.665, 0.693] | 0.950 [0.945, 0.955] | 0.223 [0.204, 0.243] | 0.718 [0.704, 0.732] |
| PsyRoBERTa | 0.879 [0.871, 0.887] | 0.375 [0.352, 0.399] | 0.704 [0.690, 0.718] | 0.933 [0.926, 0.938] | 0.293 [0.269, 0.315] | 0.728 [0.714, 0.742] |
| **Sex = Male** | | | | | | |
| LogReg | 0.671 [0.660, 0.683] | 0.602 [0.582, 0.626] | 0.691 [0.679, 0.705] | 0.508 [0.496, 0.522] | 0.800 [0.784, 0.818] | 0.723 [0.711, 0.736] |
| BERT | 0.863 [0.855, 0.872] | 0.334 [0.312, 0.355] | 0.678 [0.664, 0.691] | 0.972 [0.967, 0.976] | 0.149 [0.134, 0.164] | 0.725 [0.713, 0.738] |
| MeDa-BERT | 0.803 [0.793, 0.812] | 0.463 [0.441, 0.486] | 0.704 [0.689, 0.717] | 0.921 [0.915, 0.928] | 0.335 [0.314, 0.355] | 0.739 [0.728, 0.752] |
| RøBÆRTa | 0.891 [0.883, 0.899] | 0.318 [0.299, 0.340] | 0.695 [0.680, 0.709] | 0.945 [0.940, 0.951] | 0.249 [0.232, 0.268] | 0.736 [0.723, 0.747] |
| PsyRoBERTa | 0.836 [0.826, 0.845] | 0.456 [0.434, 0.477] | 0.716 [0.701, 0.728] | 0.905 [0.898, 0.912] | 0.365 [0.344, 0.386] | 0.742 [0.730, 0.754] |
| **Age = Above 54** | | | | | | |
| LogReg | 0.764 [0.749, 0.779] | 0.386 [0.351, 0.426] | 0.627 [0.603, 0.652] | 0.665 [0.649, 0.682] | 0.549 [0.510, 0.584] | 0.650 [0.626, 0.672] |
| BERT | 0.928 [0.918, 0.936] | 0.142 [0.117, 0.171] | 0.607 [0.585, 0.632] | 0.989 [0.985, 0.992] | 0.033 [0.021, 0.049] | 0.659 [0.635, 0.679] |
| MeDa-BERT | 0.892 [0.882, 0.902] | 0.230 [0.199, 0.266] | 0.632 [0.606, 0.656] | 0.965 [0.959, 0.971] | 0.122 [0.099, 0.150] | 0.669 [0.645, 0.688] |
| RøBÆRTa | 0.943 [0.935, 0.950] | 0.140 [0.118, 0.171] | 0.621 [0.598, 0.647] | 0.977 [0.971, 0.981] | 0.087 [0.066, 0.110] | 0.674 [0.648, 0.693] |
| PsyRoBERTa | 0.912 [0.902, 0.920] | 0.235 [0.201, 0.269] | 0.646 [0.621, 0.671] | 0.963 [0.957, 0.969] | 0.127 [0.104, 0.155] | 0.669 [0.646, 0.689] |
| **Age = 35-54** | | | | | | |
| LogReg | 0.643 [0.629, 0.659] | 0.642 [0.617, 0.667] | 0.689 [0.672, 0.706] | 0.468 [0.453, 0.483] | 0.827 [0.808, 0.845] | 0.725 [0.709, 0.739] |
| BERT | 0.854 [0.843, 0.865] | 0.365 [0.341, 0.390] | 0.685 [0.669, 0.702] | 0.960 [0.954, 0.965] | 0.184 [0.165, 0.204] | 0.730 [0.716, 0.745] |
| MeDa-BERT | 0.785 [0.773, 0.798] | 0.523 [0.498, 0.549] | 0.714 [0.699, 0.729] | 0.906 [0.897, 0.915] | 0.384 [0.362, 0.409] | 0.745 [0.730, 0.759] |
| RøBÆRTa | 0.878 [0.868, 0.888] | 0.356 [0.330, 0.380] | 0.699 [0.683, 0.716] | 0.931 [0.923, 0.938] | 0.301 [0.279, 0.324] | 0.741 [0.727, 0.756] |
| PsyRoBERTa | 0.811 [0.799, 0.822] | 0.501 [0.474, 0.526] | 0.718 [0.703, 0.734] | 0.884 [0.874, 0.894] | 0.433 [0.408, 0.457] | 0.749 [0.735, 0.763] |
| **Age = 18-24** | | | | | | |
| LogReg | 0.670 [0.657, 0.684] | 0.580 [0.557, 0.607] | 0.679 [0.662, 0.696] | 0.476 [0.461, 0.489] | 0.829 [0.806, 0.846] | 0.716 [0.701, 0.729] |
| BERT | 0.870 [0.860, 0.879] | 0.318 [0.294, 0.343] | 0.684 [0.668, 0.700] | 0.967 [0.962, 0.973] | 0.114 [0.098, 0.131] | 0.708 [0.693, 0.722] |
| MeDa-BERT | 0.797 [0.785, 0.808] | 0.476 [0.447, 0.502] | 0.697 [0.680, 0.712] | 0.921 [0.913, 0.928] | 0.297 [0.273, 0.321] | 0.733 [0.718, 0.748] |
| RøBÆRTa | 0.884 [0.875, 0.893] | 0.288 [0.264, 0.313] | 0.688 [0.672, 0.704] | 0.936 [0.928, 0.943] | 0.239 [0.218, 0.263] | 0.718 [0.703, 0.732] |
| PsyRoBERTa | 0.845 [0.834, 0.855] | 0.436 [0.410, 0.463] | 0.715 [0.700, 0.733] | 0.911 [0.902, 0.919] | 0.325 [0.301, 0.350] | 0.736 [0.720, 0.749] |
| **Age = Below 18** | | | | | | |
| LogReg | 0.825 [0.795, 0.852] | 0.304 [0.217, 0.402] | 0.607 [0.539, 0.662] | 0.689 [0.655, 0.722] | 0.661 [0.569, 0.743] | 0.736 [0.687, 0.779] |
| BERT | 0.957 [0.940, 0.971] | 0.176 [0.111, 0.267] | 0.671 [0.609, 0.723] | 0.992 [0.983, 0.996] | 0.110 [0.059, 0.190] | 0.737 [0.688, 0.781] |
| MeDa-BERT | 0.920 [0.897, 0.938] | 0.196 [0.128, 0.287] | 0.673 [0.612, 0.727] | 0.978 [0.965, 0.987] | 0.183 [0.117, 0.277] | 0.735 [0.682, 0.781] |
| RøBÆRTa | 0.953 [0.935, 0.966] | 0.206 [0.135, 0.302] | 0.665 [0.603, 0.723] | 0.977 [0.964, 0.986] | 0.183 [0.118, 0.273] | 0.738 [0.692, 0.784] |
| PsyRoBERTa | 0.954 [0.936, 0.968] | 0.127 [0.073, 0.210] | 0.677 [0.615, 0.726] | 0.961 [0.944, 0.973] | 0.193 [0.123, 0.282] | 0.743 [0.695, 0.787] |
| **Region = Capital Region** | | | | | | |
| LogReg | 0.708 [0.700, 0.717] | 0.553 [0.535, 0.573] | 0.680 [0.668, 0.692] | 0.561 [0.552, 0.572] | 0.752 [0.737, 0.768] | 0.720 [0.711, 0.732] |
| BERT | 0.897 [0.891, 0.903] | 0.274 [0.259, 0.290] | 0.673 [0.661, 0.684] | 0.978 [0.975, 0.981] | 0.110 [0.099, 0.121] | 0.722 [0.712, 0.733] |
| MeDa-BERT | 0.839 [0.832, 0.847] | 0.430 [0.414, 0.450] | 0.702 [0.690, 0.714] | 0.939 [0.935, 0.944] | 0.289 [0.274, 0.306] | 0.739 [0.729, 0.750] |
| RøBÆRTa | 0.916 [0.911, 0.921] | 0.253 [0.238, 0.269] | 0.688 [0.678, 0.700] | 0.956 [0.952, 0.960] | 0.222 [0.207, 0.238] | 0.733 [0.723, 0.744] |
| PsyRoBERTa | 0.873 [0.865, 0.879] | 0.400 [0.382, 0.422] | 0.711 [0.699, 0.722] | 0.928 [0.923, 0.933] | 0.328 [0.311, 0.346] | 0.740 [0.730, 0.751] |
| **Region = Region Zealand** | | | | | | |
| LogReg | 0.657 [0.642, 0.673] | 0.594 [0.565, 0.629] | 0.679 [0.661, 0.699] | 0.466 [0.449, 0.481] | 0.835 [0.810, 0.857] | 0.714 [0.697, 0.732] |
| BERT | 0.852 [0.840, 0.864] | 0.383 [0.351, 0.415] | 0.696 [0.677, 0.716] | 0.956 [0.949, 0.962] | 0.182 [0.160, 0.207] | 0.709 [0.691, 0.727] |
| MeDa-BERT | 0.788 [0.774, 0.802] | 0.485 [0.451, 0.516] | 0.696 [0.677, 0.715] | 0.908 [0.898, 0.917] | 0.329 [0.301, 0.360] | 0.722 [0.704, 0.739] |
| RøBÆRTa | 0.860 [0.849, 0.872] | 0.386 [0.354, 0.418] | 0.693 [0.674, 0.713] | 0.925 [0.916, 0.934] | 0.277 [0.248, 0.307] | 0.717 [0.699, 0.735] |
| PsyRoBERTa | 0.816 [0.804, 0.830] | 0.473 [0.441, 0.509] | 0.716 [0.697, 0.735] | 0.893 [0.884, 0.902] | 0.344 [0.317, 0.377] | 0.727 [0.709, 0.744] |

True Positive Rate (TPR), True Negative Rate (TNR) and Area Under the ROC Curve (AUC) across demographic subgroups.

### Figure S3. Diagnosis fairness

| Model | TPR | TNR | AUROC | TPR | TNR | AUROC |
|---|---|---|---|---|---|---|
| | Data = discharge summaries | | | Data = all admission notes (deduplicated) | | |
| **Diagnosis = Anxiety/OCD** | | | | | | |
| LogReg | 0.766 [0.710, 0.808] | 0.355 [0.241, 0.486] | 0.562 [0.483, 0.643] | 0.664 [0.606, 0.711] | 0.530 [0.398, 0.641] | 0.601 [0.514, 0.671] |
| BERT | 0.942 [0.912, 0.966] | 0.065 [0.017, 0.153] | 0.535 [0.459, 0.615] | 0.988 [0.971, 0.997] | 0.0 | 0.618 [0.538, 0.683] |
| MeDa-BERT | 0.926 [0.893, 0.952] | 0.129 [0.058, 0.240] | 0.574 [0.490, 0.652] | 0.991 [0.975, 0.997] | 0.015 [0.000, 0.085] | 0.626 [0.538, 0.693] |
| RøBÆRTa | 0.965 [0.937, 0.981] | 0.032 [0.000, 0.104] | 0.561 [0.488, 0.635] | 0.981 [0.961, 0.991] | 0.015 [0.000, 0.085] | 0.645 [0.565, 0.705] |
| PsyRoBERTa | 0.942 [0.908, 0.964] | 0.113 [0.050, 0.209] | 0.604 [0.523, 0.674] | 0.978 [0.957, 0.991] | 0.061 [0.016, 0.150] | 0.662 [0.583, 0.725] |
| **Diagnosis = Personality Disorder** | | | | | | |
| LogReg | 0.573 [0.525, 0.616] | 0.720 [0.656, 0.781] | 0.706 [0.663, 0.748] | 0.295 [0.257, 0.336] | 0.919 [0.878, 0.952] | 0.734 [0.697, 0.771] |
| BERT | 0.875 [0.841, 0.901] | 0.259 [0.196, 0.324] | 0.663 [0.620, 0.703] | 0.964 [0.946, 0.978] | 0.158 [0.111, 0.210] | 0.735 [0.694, 0.775] |
| MeDa-BERT | 0.734 [0.690, 0.771] | 0.614 [0.548, 0.684] | 0.710 [0.663, 0.752] | 0.886 [0.859, 0.911] | 0.469 [0.405, 0.540] | 0.765 [0.727, 0.803] |
| RøBÆRTa | 0.884 [0.853, 0.909] | 0.270 [0.209, 0.333] | 0.650 [0.605, 0.698] | 0.906 [0.880, 0.929] | 0.426 [0.358, 0.500] | 0.756 [0.718, 0.794] |
| PsyRoBERTa | 0.784 [0.742, 0.818] | 0.587 [0.520, 0.658] | 0.724 [0.676, 0.769] | 0.880 [0.852, 0.907] | 0.531 [0.462, 0.601] | 0.771 [0.732, 0.808] |
| **Diagnosis = Bipolar/Manic** | | | | | | |
| LogReg | 0.750 [0.720, 0.779] | 0.403 [0.332, 0.467] | 0.621 [0.574, 0.662] | 0.735 [0.703, 0.762] | 0.538 [0.470, 0.607] | 0.693 [0.651, 0.730] |
| BERT | 0.950 [0.935, 0.964] | 0.149 [0.101, 0.206] | 0.615 [0.568, 0.659] | 0.998 [0.992, 1.000] | 0.014 [0.004, 0.037] | 0.714 [0.675, 0.747] |
| MeDa-BERT | 0.911 [0.889, 0.928] | 0.264 [0.198, 0.323] | 0.655 [0.608, 0.698] | 0.984 [0.973, 0.991] | 0.077 [0.047, 0.116] | 0.726 [0.685, 0.758] |
| RøBÆRTa | 0.947 [0.931, 0.961] | 0.139 [0.095, 0.189] | 0.661 [0.618, 0.700] | 0.987 [0.976, 0.992] | 0.059 [0.032, 0.097] | 0.723 [0.683, 0.756] |
| PsyRoBERTa | 0.934 [0.915, 0.950] | 0.234 [0.181, 0.295] | 0.684 [0.640, 0.722] | 0.978 [0.965, 0.985] | 0.081 [0.052, 0.125] | 0.712 [0.671, 0.747] |
| **Diagnosis = Depression** | | | | | | |
| LogReg | 0.861 [0.844, 0.880] | 0.281 [0.217, 0.338] | 0.617 [0.571, 0.657] | 0.863 [0.845, 0.88] | 0.311 [0.253, 0.376] | 0.659 [0.618, 0.700] |
| BERT | 0.988 [0.981, 0.992] | 0.057 [0.029, 0.098] | 0.593 [0.547, 0.633] | 1.0 | 0.0 | 0.646 [0.606, 0.686] |
| MeDa-BERT | 0.976 [0.967, 0.983] | 0.081 [0.049, 0.122] | 0.618 [0.576, 0.659] | 0.998 [0.995, 0.999] | 0.014 [0.004, 0.040] | 0.654 [0.614, 0.693] |
| RøBÆRTa | 0.991 [0.985, 0.995] | 0.024 [0.009, 0.051] | 0.633 [0.589, 0.672] | 0.998 [0.995, 0.999] | 0.005 [0.000, 0.023] | 0.656 [0.618, 0.697] |
| PsyRoBERTa | 0.989 [0.982, 0.993] | 0.071 [0.040, 0.111] | 0.617 [0.572, 0.658] | 0.997 [0.993, 0.999] | 0.005 [0.000, 0.023] | 0.660 [0.622, 0.697] |
| **Diagnosis = Other Psychosis** | | | | | | |
| LogReg | 0.730 [0.706, 0.754] | 0.454 [0.401, 0.501] | 0.641 [0.610, 0.672] | 0.632 [0.609, 0.657] | 0.696 [0.648, 0.739] | 0.718 [0.688, 0.745] |
| BERT | 0.925 [0.911, 0.938] | 0.260 [0.218, 0.307] | 0.652 [0.621, 0.685] | 0.995 [0.990, 0.998] | 0.049 [0.030, 0.073] | 0.716 [0.687, 0.744] |
| MeDa-BERT | 0.872 [0.852, 0.888] | 0.329 [0.283, 0.377] | 0.671 [0.639, 0.702] | 0.974 [0.964, 0.981] | 0.199 [0.163, 0.239] | 0.738 [0.708, 0.765] |
| RøBÆRTa | 0.937 [0.921, 0.949] | 0.228 [0.188, 0.274] | 0.659 [0.628, 0.691] | 0.983 [0.975, 0.989] | 0.141 [0.107, 0.176] | 0.724 [0.697, 0.752] |
| PsyRoBERTa | 0.907 [0.890, 0.922] | 0.294 [0.251, 0.343] | 0.670 [0.637, 0.701] | 0.973 [0.964, 0.981] | 0.199 [0.161, 0.238] | 0.731 [0.702, 0.758] |
| **Diagnosis = SUD** | | | | | | |
| LogReg | 0.564 [0.540, 0.586] | 0.580 [0.539, 0.618] | 0.609 [0.584, 0.635] | 0.242 [0.223, 0.260] | 0.887 [0.860, 0.910] | 0.614 [0.590, 0.640] |
| BERT | 0.834 [0.818, 0.852] | 0.285 [0.251, 0.326] | 0.616 [0.591, 0.642] | 0.962 [0.953, 0.970] | 0.077 [0.057, 0.102] | 0.622 [0.597, 0.649] |
| MeDa-BERT | 0.735 [0.715, 0.755] | 0.441 [0.401, 0.480] | 0.634 [0.607, 0.660] | 0.878 [0.865, 0.892] | 0.257 [0.225, 0.295] | 0.657 [0.633, 0.683] |
| RøBÆRTa | 0.875 [0.860, 0.891] | 0.226 [0.194, 0.266] | 0.620 [0.594, 0.647] | 0.915 [0.902, 0.927] | 0.169 [0.142, 0.202] | 0.648 [0.623, 0.672] |
| PsyRoBERTa | 0.778 [0.758, 0.797] | 0.392 [0.353, 0.432] | 0.640 [0.614, 0.666] | 0.848 [0.831, 0.864] | 0.328 [0.294, 0.366] | 0.664 [0.641, 0.690] |
| **Diagnosis = Schizophrenia** | | | | | | |
| LogReg | 0.584 [0.567, 0.602] | 0.683 [0.657, 0.709] | 0.688 [0.669, 0.704] | 0.385 [0.368, 0.401] | 0.889 [0.871, 0.907] | 0.734 [0.719, 0.750] |
| BERT | 0.774 [0.760, 0.789] | 0.447 [0.420, 0.474] | 0.673 [0.657, 0.691] | 0.937 [0.928, 0.945] | 0.219 [0.198, 0.243] | 0.730 [0.713, 0.745] |
| MeDa-BERT | 0.693 [0.675, 0.708] | 0.598 [0.572, 0.624] | 0.699 [0.682, 0.716] | 0.865 [0.853, 0.877] | 0.434 [0.408, 0.462] | 0.746 [0.730, 0.761] |
| RøBÆRTa | 0.806 [0.790, 0.818] | 0.424 [0.398, 0.452] | 0.679 [0.663, 0.696] | 0.899 [0.888, 0.909] | 0.367 [0.340, 0.392] | 0.747 [0.731, 0.763] |
| PsyRoBERTa | 0.741 [0.723, 0.756] | 0.584 [0.559, 0.613] | 0.719 [0.703, 0.737] | 0.848 [0.835, 0.861] | 0.480 [0.453, 0.508] | 0.750 [0.734, 0.765] |
| **Diagnosis = Other** | | | | | | |
| LogReg | 0.778 [0.764, 0.793] | 0.510 [0.472, 0.549] | 0.700 [0.677, 0.720] | 0.625 [0.609, 0.641] | 0.708 [0.673, 0.739] | 0.724 [0.702, 0.745] |
| BERT | 0.933 [0.924, 0.941] | 0.240 [0.208, 0.273] | 0.701 [0.678, 0.722] | 0.982 [0.976, 0.985] | 0.135 [0.111, 0.160] | 0.727 [0.702, 0.745] |
| MeDa-BERT | 0.894 [0.883, 0.905] | 0.386 [0.352, 0.426] | 0.714 [0.691, 0.736] | 0.962 [0.955, 0.968] | 0.284 [0.253, 0.319] | 0.733 [0.710, 0.753] |
| RøBÆRTa | 0.935 [0.925, 0.942] | 0.288 [0.255, 0.323] | 0.717 [0.694, 0.738] | 0.967 [0.961, 0.973] | 0.205 [0.175, 0.234] | 0.723 [0.701, 0.743] |
| PsyRoBERTa | 0.914 [0.904, 0.923] | 0.363 [0.329, 0.400] | 0.725 [0.704, 0.747] | 0.952 [0.943, 0.958] | 0.286 [0.252, 0.319] | 0.740 [0.718, 0.760] |

True Positive Rate (TPR), True Negative Rate (TNR) and Area Under the ROC Curve (AUC) across psychiatric diagnosis subgroups.

### Figure S4. Min-max difference in performance across subgroups

| Attribute | AUROC Min-Max difference | |
|---|---|---|
| Model | Data = dis. sum. | Data = all |
| **Sex** | | |
| LogReg | 0.026 | 0.012 |
| BERT | 0.002 | 0.017 |
| MeDa-BERT | 0.011 | 0.012 |
| RøBÆRTa | 0.017 | 0.018 |
| PsyRoBERTa | 0.012 | 0.014 |
| **Age** | | |
| LogReg | 0.082 | 0.086 |
| BERT | 0.078 | 0.078 |
| MeDa-BERT | 0.082 | 0.076 |
| RøBÆRTa | 0.078 | 0.068 |
| PsyRoBERTa | 0.071 | 0.079 |
| **Region** | | |
| LogReg | 0.001 | 0.006 |
| BERT | 0.023 | 0.013 |
| MeDa-BERT | 0.006 | 0.017 |
| RøBÆRTa | 0.004 | 0.016 |
| PsyRoBERTa | 0.005 | 0.013 |
| **Diagnosis** | | |
| LogReg | 0.144 | 0.133 |
| BERT | 0.166 | 0.117 |
| MeDa-BERT | 0.140 | 0.139 |
| RøBÆRTa | 0.156 | 0.112 |
| PsyRoBERTa | 0.122 | 0.111 |

Differences in the minimum and maximum AUROC across subgroups within the attributes sex, age, region and psychiatric diagnosis. A smaller difference entails greater fairness by performance parity.

#### Figure S5. Diagnosis fairness analysis of PsyRoBERTa finetuned on all admission notes

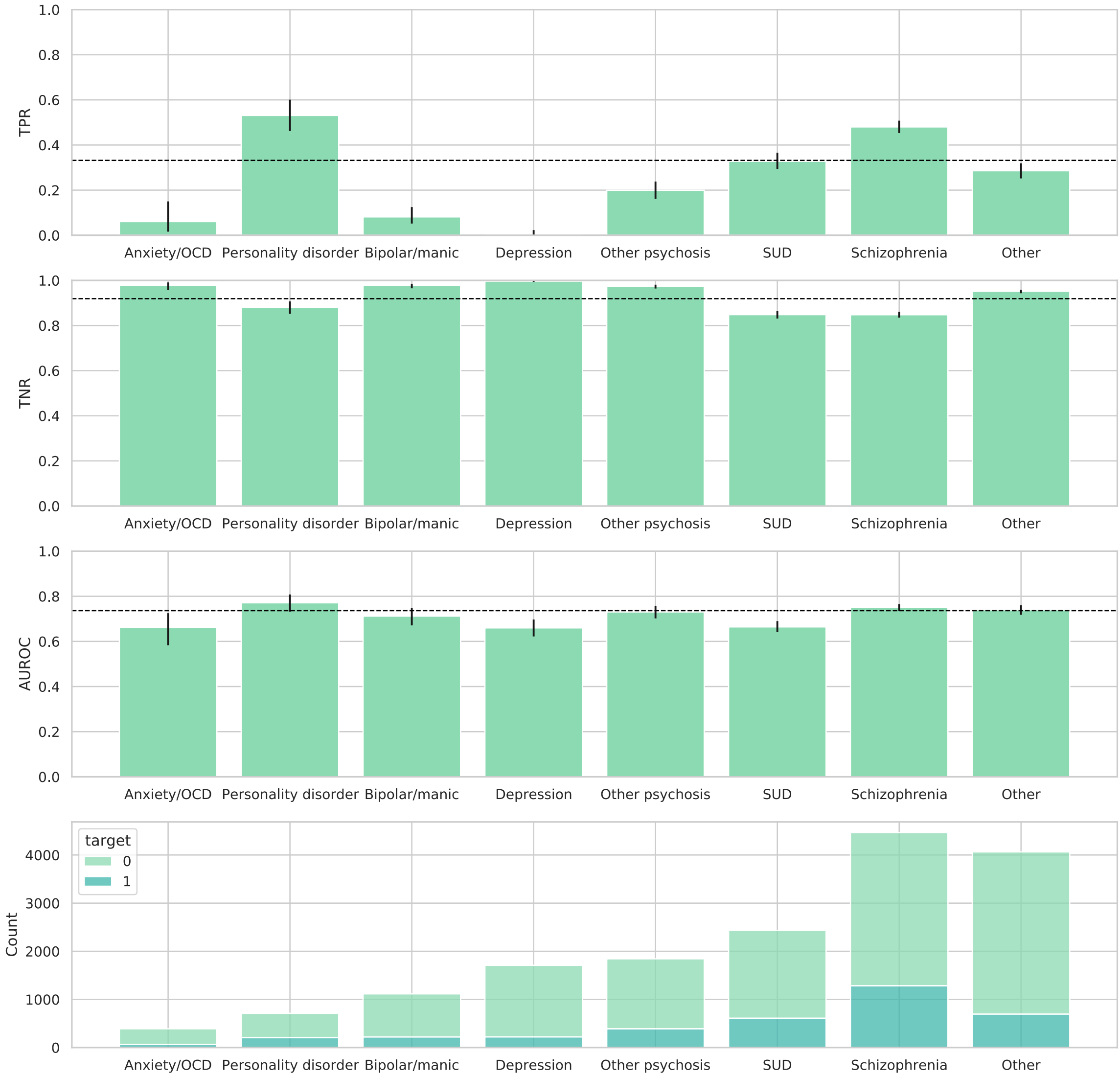


Fairness was evaluated through the lens of equalized odds (equality in true positive and true negative rates) and performance parity.[2,3] The dotted lines indicate overall rates and performance on the test set while the bars show the rates and performance for subgroups. Subgroups were defined as the most severe diagnosis at the time of admission. The bottom bar chart shows the count of admissions within the subgroups and targets being whether the next admission was acute (1) and not acute (0).

#### Figure S6. Demographics fairness analysis of PsyRoBERTa finetuned on all admission notes

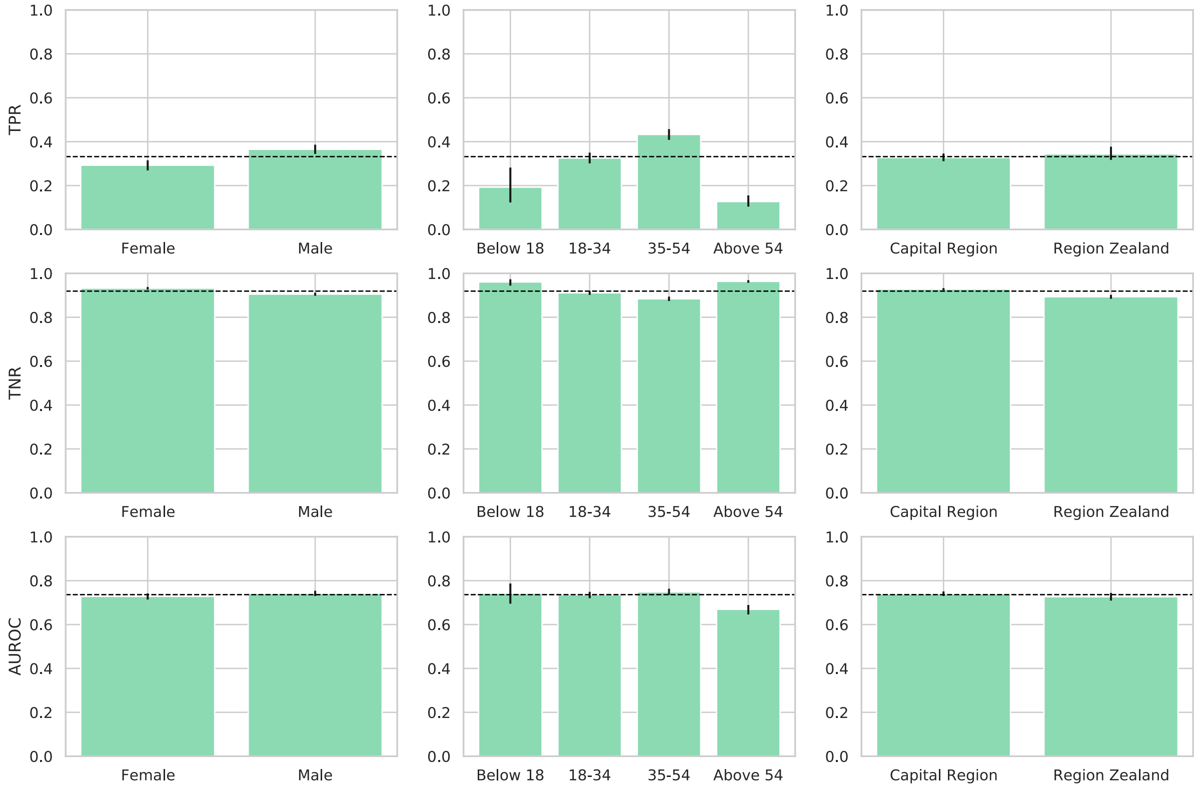


Fairness across sex, age groups, and region was evaluated through the lens of equalized odds (equality in true positive and true negative rates) and performance parity.[2,3] The dotted lines indicate the rates and performance on the test set, and the bars indicate the subgroup rates and performance within the test set.

#### Figure S7. Diagnosis fairness analysis of PsyRoBERTa finetuned on discharge summaries

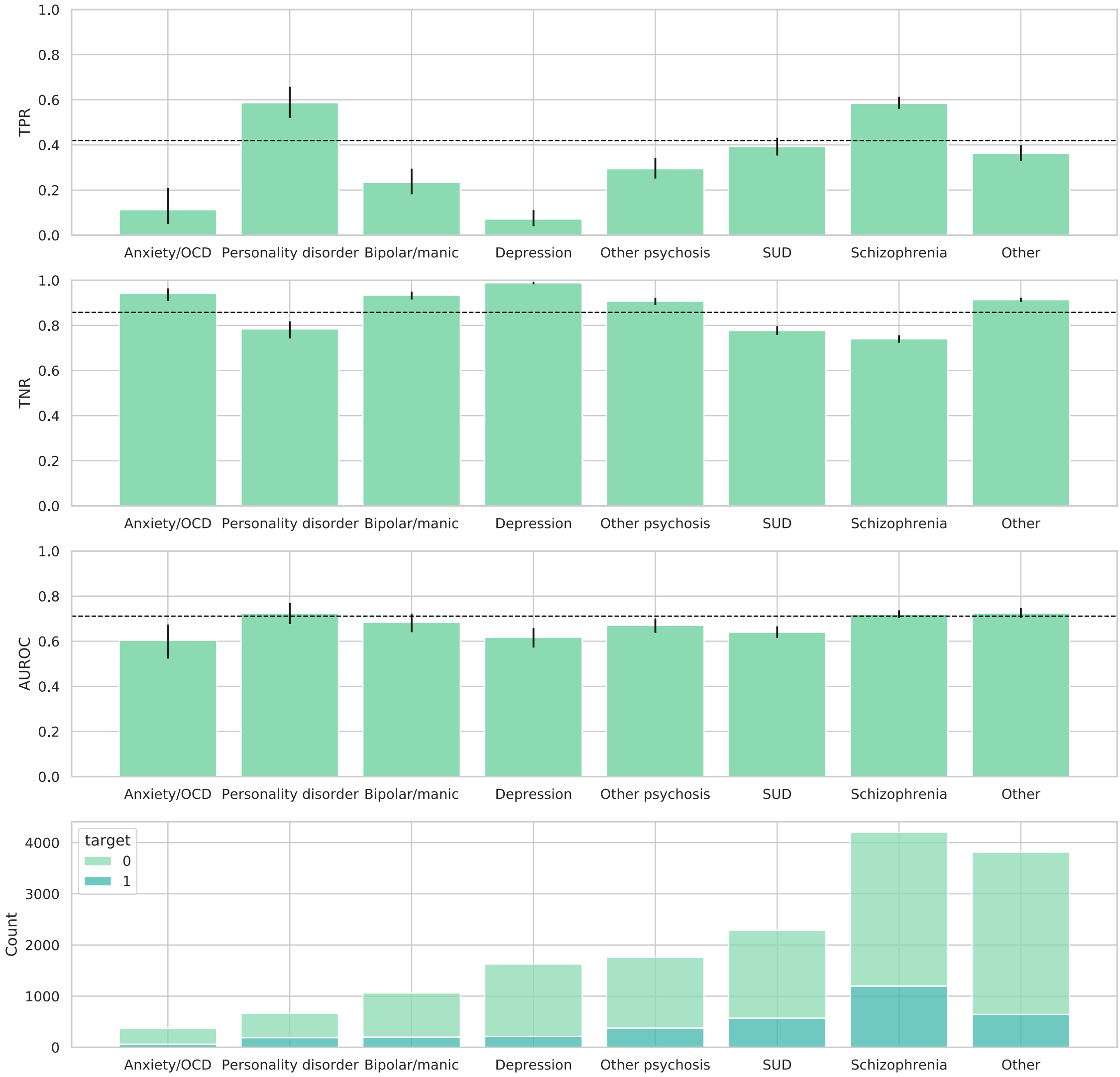


Fairness was evaluated through the lens of equalized odds (equality in true positive and true negative rates) and performance parity.[2,3] The dotted lines indicate overall rates and performance on the test set while the bars show the rates and performance for subgroups. Subgroups were defined as the most severe diagnosis at the time of admission. The bottom bar chart shows the count of admissions within the subgroups and targets being whether the next admission was acute (1) and not acute (0).

#### Figure S8. Demographics fairness analysis of PsyRoBERTa finetuned on discharge summaries

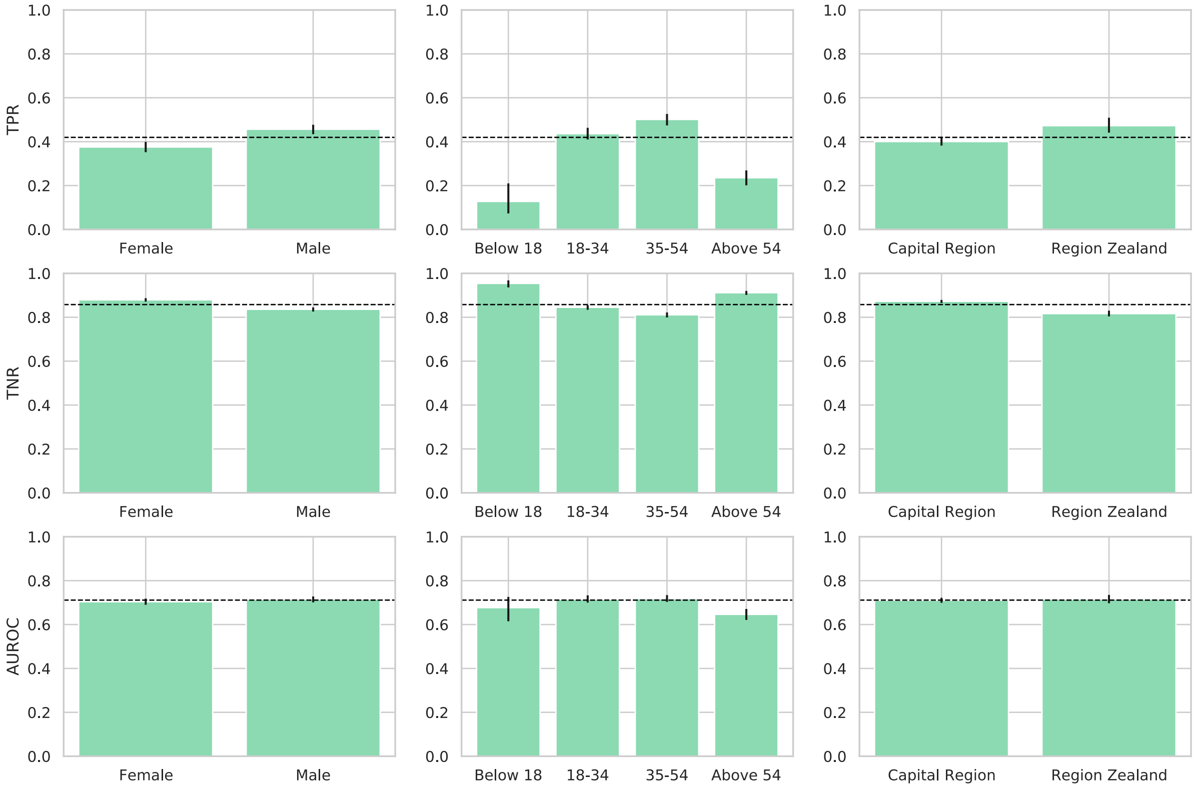


Fairness across sex, age groups, and region was evaluated through the lens of equalized odds (equality in true positive and true negative rates) and performance parity.[2,3] The dotted lines indicate the rates and performance on the test set, and the bars indicate the subgroup rates and performance within the test set.

#### Figure S9. Impact of increasing pretraining data on the downstream task of predicting acute readmissions from discharge summaries

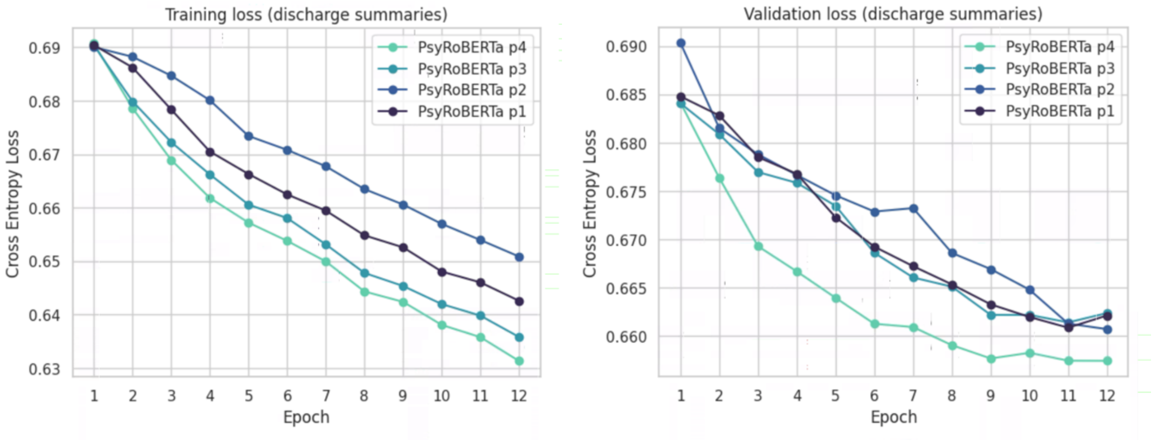


PsyRoBERTa p1-4 denotes the checkpoints of PsyRoBERTa through incremental pretraining with the four data parts summarized in **Figure 2B** and here finetuned with discharge summaries. PsyRoBERTa p4 is the model pretrained on all data parts and used in the following experiments.

#### Figure S10. Language model data requirement analysis

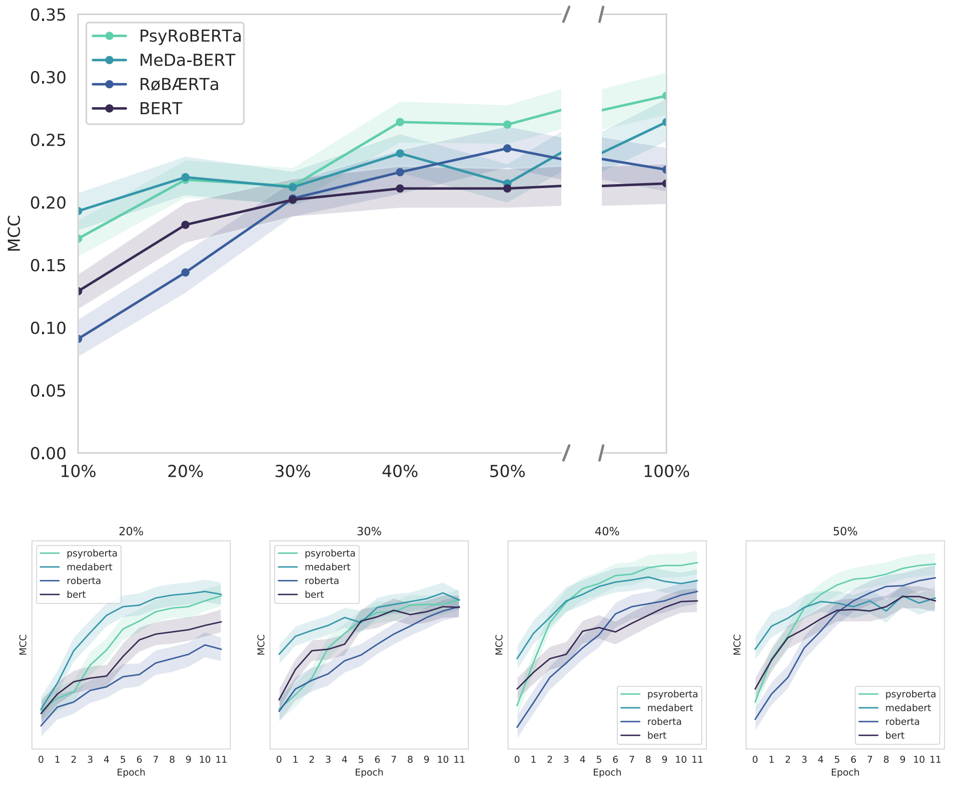


Language models finetuned on 10%, 20%, 30%, 40% and 50% of the discharge summary training data. The top plot shows the MCC of the last epoch (12) for each model given the percentage of training data, and it shows that PsyRoBERTa and MeDa-BERT had better onsets in performance. The bottom plot shows the per-epoch performance for each percentage of training data and shows that PsyRoBERTa and MeDa-BERT generally improved faster.

### Figure S11. Top 25 most highly weighted bigrams and trigrams by Attention Rollout score

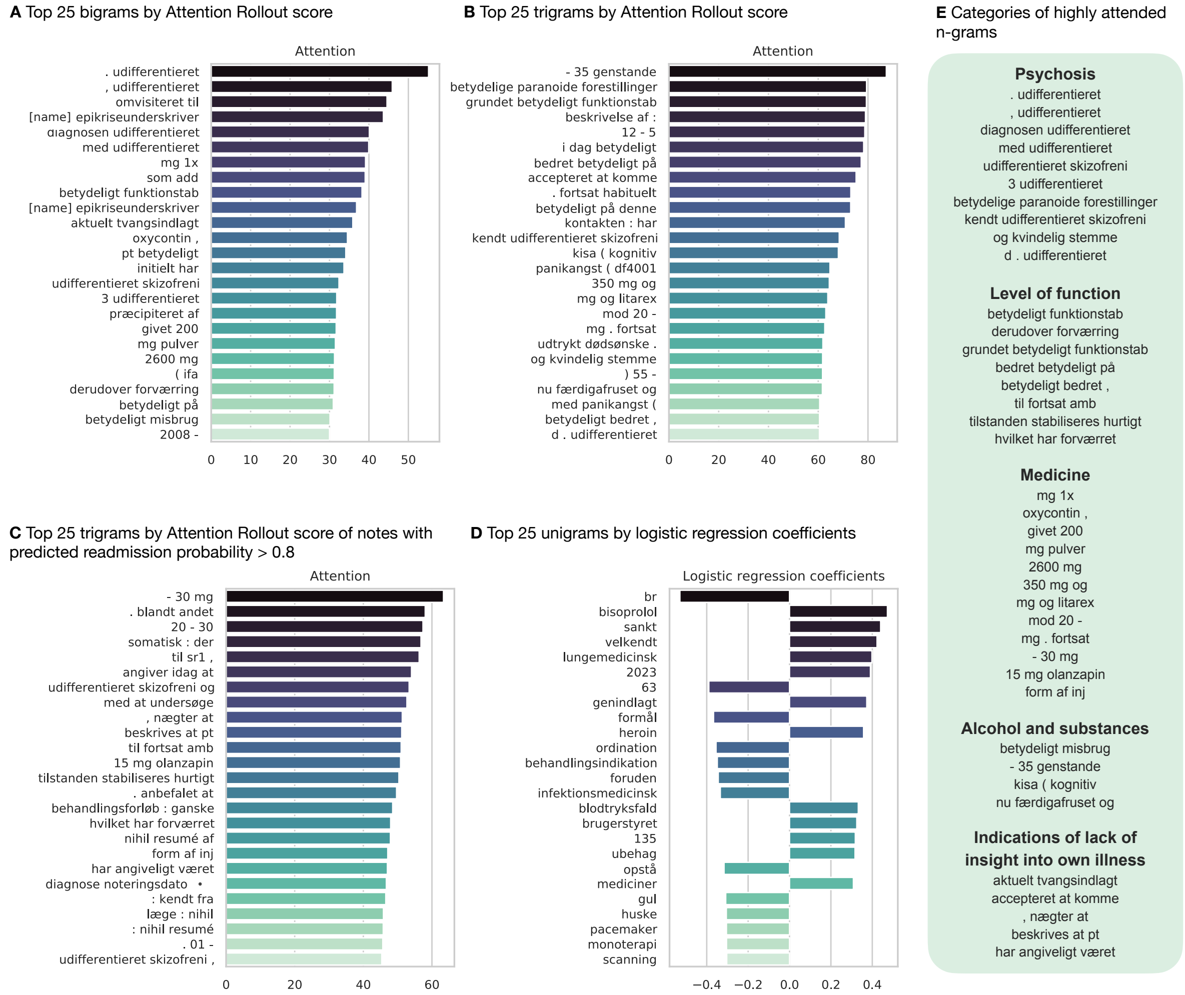


This is the Danish (original) version of **Figure 4**.

**Figure S12. Danish and English translations of the most highly weighted ngrams with their mean and max AR scores**

**A**

| Unigrams | | | | | |
|---|---|---|---|---|---|
| Mean | | | Max | | |
| AR | EN | DA (original) | AR | EN | DA (original) |
| 18.34 | and | och | 31.01 | undifferentiated | udifferentieret |
| 13.07 | significantly | betydeligt | 18.65 | note date | noteringsdato |
| 12.47 | belongs to / is part of | tilhører | 18.34 | and | och |
| 11.80 | swedish | svensk | 13.35 | 21735 | 21735 |
| 11.68 | plenty | rigeligt | 13.07 | significantly | betydeligt |
| 10.25 | plenty | masser | 12.80 | psychoeducational | psykoedukative |
| 9.96 | significant | betydelige | 12.49 | belongs to / is part of | tilhører |
| 9.54 | refugees | flygtninge | 11.80 | swedish | svensk |
| 9.17 | undifferentiated | udifferentieret | 11.68 | plenty | rigeligt |
| 8.96 | winter | vinter | 11.63 | discharge summary | epikriseunderskriver |
| 8.46 | additionally | derudover | 10.75 | reflected | reflekteret |
| 7.96 | professor | professor | 10.71 | brings | medbringer |
| 7.78 | 21735 | 21735 | 10.43 | 90mg | 90mg |
| 7.77 | explains/tells | forteller | 10.41 | alcoholism | alkoholisme |
| 7.76 | maintaining | vedligeholdelse | 10.25 | plenty | masser |
| 7.63 | approximately | omtrent | 10.11 | respirator treatment | respiratorbehandling |
| 7.55 | powder | pulver | 10.08 | df312 | df312 |
| 7.48 | note date | noteringsdato | 9.96 | significant | betydelige |
| 7.26 | iraq | irak | 9.66 | opt-out | fravalg |
| 6.99 | often | hyppigt | 9.62 | refugees | flygtninge |
| 6.92 | looked/gazed | kiggede | 9.47 | extended-release granules | depotgranulat |
| 6.91 | naturally | selvfølgelig | 9.11 | serdolect | serdolect |
| 6.77 | 90mg | 90mg | 9.03 | [name] | [name] |
| 6.74 | total | totalt | 8.96 | winter | vinter |
| 6.73 | usa | usa | 8.95 | co-patient | medptt |

**B**

| Bigrams | | | | | |
|---|---|---|---|---|---|
| Mean | | | Max | | |
| AR | EN | DA (original) | AR | EN | DA (original) |
| 41.92 | reassigned to | omvisiteret til | 55.12 | . undifferentiated | . udifferentieret |
| 39.05 | like add | som add | 45.91 | , undifferentiated | , udifferentieret |
| 36.69 | significant loss of function | betydeligt funktionstab | 44.58 | reassigned to | omvisiteret til |
| 34.12 | pt significantly | pt betydeligt | 43.67 | [name] discharge summary signatory | [name] epikriseunderskriver |
| 33.66 | currently involuntarily admitted | aktuelt tvangsindlagt | 40.10 | the diagnosis undifferentiated | diagnosen udifferentieret |
| 32.49 | has initially | initielt har | 39.95 | with undifferentiated | med udifferentieret |
| 31.5 | mg powder | mg pulver | 39.14 | mg 1x | mg 1x |
| 31.23 | given 200 | givet 200 | 39.05 | like add | som add |
| 31.21 | ( in the form of | ( ifa | 38.24 | significant loss of function | betydeligt funktionstab |
| 30.96 | significantly on | betydeligt på | 36.86 | [name] discharge summary signatory | [name] epikriseunderskriver |
| 30.10 | significant abuse | betydeligt misbrug | 35.91 | currently involuntarily admitted | aktuelt tvangsindlagt |
| 30.01 | -2008 | -2008 | 34.55 | oxycontin , | oxycontin , |
| 29.73 | additionally worsening | derudover forværring | 34.13 | pt significantly | pt betydeligt |
| 29.31 | not global | ej globalt | 33.67 | has initially | initielt har |
| 28.97 | precipitated by | præcipiteret af | 32.40 | undifferentiated schizophrenia | udifferentieret skizofreni |
| 28.67 | improved significantly | bedret betydeligt | 31.84 | 3 undifferentiated | 3 udifferentieret |
| 28.37 | adopted by | adopteret fra | 31.79 | precipitated by | præcipiteret af |
| 27.59 | otherwise own | ellers egen | 31.69 | given 200 | givet 200 |
| 27.28 | accepted to | accepteret at | 31.5 | mg powder | mg pulver |
| 27.19 | considered to | overvejet at | 31.27 | 2600 mg | 2600 mg |
| 27.15 | last winter | sidste vinter | 31.21 | ( in the form of | ( ifa |
| 26.73 | has had significant | haft betydelige | 31.17 | additionally worsening | derudover forværring |
| 26.55 | powder and | pulver og | 30.96 | significantly on | betydeligt på |
| 26.46 | a danish | en dansk | 30.13 | significant abuse | betydeligt misbrug |
| 26.45 | 15 months | 15 mdr | 30.01 | -2008 | -2008 |

**C**

| Trigrams | | | | | |
|---|---|---|---|---|---|
| Mean | | | Max | | |
| AR | EN | DA (original) | AR | EN | DA (original) |
| 87.37 | - 35 units | - 35 genstande | 87.37 | - 35 units | - 35 genstande |
| 78.99 | report by : | beskrivelse af : | 79.43 | significant paranoid delusions | betydelige paranoide forestillinger |
| 78.72 | 12 – 5 | 12 - 5 | 79.36 | because of significant loss of function | grundet betydeligt funktionstab |
| 78.27 | today significantly | i dag betydeligt | 78.99 | report by : | beskrivelse af : |
| 76.49 | because of significant loss of function | grundet betydeligt funktionstab | 78.72 | 12 – 5 | 12 - 5 |
| 75.79 | significant paranoid delusions | betydelige paranoide forestillinger | 78.27 | today significantly | i dag betydeligt |
| 74.79 | improved significantly on | bedret betydeligt på | 77.28 | improved significantly on | bedret betydeligt på |
| 73.26 | accepted to come | accepteret at komme | 75.23 | accepted to come | accepteret at komme |
| 73.06 | significantly on this | betydeligt på denne | 73.12 | . still habitual | . fortsat habituelt |
| 70.92 | the contact : has | kontakten : har | 73.06 | significantly on this | betydeligt på denne |
| 64.45 | 350 mg and | 350 mg og | 70.92 | the contact : has | kontakten : har |
| 63.06 | towards 20 – | mod 20 - | 68.44 | known undifferentiated schizophrenia | kendt udifferentieret skizofreni |
| 61.73 | ) 55 – | ) 55 - | 68.10 | kisa ( cognitive | kisa ( kognitiv |
| 60.80 | mg and litarex | mg og litarex | 64.73 | panic disorder ( df4001 | panikangst ( df4001 |
| 60.34 | because of 4 - | grundet 4 - | 64.45 | 350 mg and | 350 mg og |
| 60.04 | adjusted/corrected themself significantly | rettet sig betydeligt | 63.86 | mg and litarex | mg og litarex |
| 59.74 | panic disorder ( df4001 | panikangst ( df4001 | 63.06 | towards 20 – | mod 20 - |
| 59.59 | the contact : 31 | kontakten : 31 | 62.64 | mg . continued | mg . fortsat |
| 58.94 | and female voice | og kvindelig stemme | 61.88 | expressed death wish | udtrykt dødsønske . |
| 58.86 | now sobered and | nu færdigafruset og | 61.77 | and female voice | og kvindelig stemme |
| 58.74 | with panic disorder | med panikangst ( | 61.73 | ) 55 - | ) 55 - |
| 58.68 | reports that the police | fortæller at politiet | 61.67 | now sobered and | nu færdigafruset og |
| 58.64 | suicidal thoughts report by | selvmordstanker beskrivelse af | 60.59 | with panic disorder ( | med panikangst ( |
| 58.26 | an appoinment as soon as possible | en tid snarest | 60.51 | significantly improved , | betydeligt bedret , |
| 57.99 | significantly improved , | betydeligt bedret , | 60.49 | d . undifferentiated | d . udifferentieret |

Top 25 **A**. unigrams, **B**. bigrams, and **C**. trigrams with highest **A**ttention **R**ollout attribution score of the 10% most frequent n-grams globally.

### Figure S13. Danish and English translations of the most highly weighted ngrams with their mean and max AR scores (high risk instances)

**A**

| Unigrams | | | | | |
|---|---|---|---|---|---|
| Mean | | | Max | | |
| AR | EN | DA (original) | AR | EN | DA (original) |
| 37.69 | belongs to | tilhører | 38.42 | undifferentiated | udifferentieret |
| 25.54 | nice/lovely | dejligt | 37.69 | belongs to | tilhører |
| 20.67 | significant | betydelige | 35.57 | note date | noteringsdato |
| 16.37 | even | endda | 25.99 | brought | medbragt |
| 14.98 | brought | medbragt | 25.54 | nice/lovely | dejligt |
| 14.42 | total | totalt | 20.67 | significant | betydelige |
| 14.35 | _ | _ | 19.63 | co-patient | medptt |
| 14.28 | points (on Hamilton rating scale) | pkt | 17.80 | contacted | kontaktede |
| 14.17 | fast | hurtige | 16.37 | even | endda |
| 13.86 | note date | noteringsdato | 16.37 | oxycontin | oxycontin |
| 13.45 | sex | sex | 16.12 | smoke | røg |
| 12.90 | global | globalt | 15.57 | 150mg | 150mg |
| 11.61 | whereof | hvoraf | 14.84 | normalized | normaliseret |
| 11.14 | in return | gengæld | 14.42 | total | totalt |
| 11.13 | additionally | derudover | 14.35 | _ | _ |
| 11.05 | undifferentiated | udifferentieret | 14.28 | points (on Hamilton rating scale) | pkt |
| 10.89 | 1200 | 1200 | 14.17 | fast | hurtige |
| 10.83 | contacted | kontaktede | 13.45 | sex | sex |
| 10.29 | guidance/advice/consultation | rådgivning | 13.34 | present/in attendance | tilsted |
| 10.25 | improvement | forbedring | 12.97 | administratively | administrativt |
| 9.90 | powder | pulver | 12.93 | global | globalt |
| 9.75 | 150mg | 150mg | 12.63 | §108 | §108 |
| 9.61 | significantly | betydeligt | 12.60 | suspicious | suspekt |
| 9.48 | some/certain (things) | visse | 11.61 | whereof | hvoraf |
| 9.32 | smoke | røg | 11.60 | The correctional service in freedom | kif |

**B**

| Bigrams | | | | | |
|---|---|---|---|---|---|
| Mean | | | Max | | |
| AR | EN | DA (original) | AR | EN | DA (original) |
| 57.83 | confronted with | konfronteret med | 59.41 | confronted with | konfronteret med |
| 52.74 | . among | . blandt | 53.37 | improved significantly | bedret betydeligt |
| 51.25 | improved significantly | bedret betydeligt | 52.74 | . among | . blandt |
| 49.08 | expressed that | udtalt at | 51.73 | expressed that | udtalt at |
| 47.87 | reports today | angiver idag | 47.89 | reports today | angiver idag |
| 46.65 | : quite | : ganske | 46.65 | : quite | : ganske |
| 44.34 | follow-up for | opfølgning til | 44.34 | follow-up for | opfølgning til |
| 41.71 | considered to | overvejet at | 43.04 | none/missing summary | nihil resumé |
| 40.32 | accepted to | accepteret at | 42.64 | considered to | overvejet at |
| 39.96 | superficial and | overfladisk og | 42.45 | diagnosis note date | diagnose noteringsdato |
| 38.42 | with significant | med betydelige | 41.39 | accepted to | accepteret at |
| 38.33 | caused by a | skyldes en | 41.35 | superficial and | overfladisk og |
| 37.77 | at parents | hos forældre | 40.21 | destabilized by | destabiliseret af |
| 37.61 | with respect to stabilization | mht stabilisering | 39.83 | diagnosis note date • | noteringsdato • |
| 37.28 | destabilized by | destabiliseret af | 39.45 | undifferentiated schizophrenia | udifferentieret skizofreni |
| 37.01 | ) . | ) . | 39.33 | injection by/of/with | injektion af |
| 36.88 | will firstly | først vil | 39.28 | with respect to stabilization | mht stabilisering |
| 36.38 | upset and | oprevet og | 38.42 | with significant | med betydelige |
| 35.98 | - c | - c | 38.33 | caused by a | skyldes en |
| 34.83 | significantly below/during | betydeligt under | 37.77 | at parents | hos forældre |
| 34.52 | voice and | stemme og | 37.65 | upset and | oprevet og |
| 34.51 | -24 | -24 | 37.01 | ) . | ) . |
| 33.64 | quite stable | ganske stabil | 36.88 | will firstly | først vil |
| 33.57 | of/by injection | af inj | 36.62 | with undifferentiated | med udifferentieret |
| 32.75 | with significant | med betydelig | 35.98 | - c | - c |

**C**

| Trigrams | | | | | |
|---|---|---|---|---|---|
| Mean | | | Max | | |
| AR | EN | DA (original) | AR | EN | DA (original) |
| 63.37 | - 30 mg | - 30 mg | 63.37 | - 30 mg | - 30 mg |
| 58.04 | . such as / among other things | . blandt andet | 58.04 | . such as / among other things | . blandt andet |
| 57.38 | 20 - 30 | 20 - 30 | 57.39 | 20 - 30 | 20 - 30 |
| 54.03 | reports today that | angiver idag at | 56.81 | somatic : there | somatisk : der |
| 52.70 | with investigating/examining | med at undersøge | 56.28 | to sr1 (suicide risk 1) | til sr1 , |
| 51.39 | somatic : there | somatisk : der | 54.05 | reports today that | angiver idag at |
| 51.19 | reported/described that pt | beskrives at pt | 53.35 | undifferentiated schizophrenia and | udifferentieret skizofreni og |
| 51.03 | for continued ambulatory | til fortsat amb | 52.71 | with investigating/examining | med at undersøge |
| 49.29 | . recommended that | . anbefalet at | 51.36 | , refuse to | , nægter at |
| 49.04 | , refuse to | , nægter at | 51.20 | reported/described that pt | beskrives at pt |
| 48.33 | course of treatment : quite | behandlingsforløb : ganske | 51.03 | for continued ambulatory | til fortsat amb |
| 47.13 | in the form of injectable | form af inj | 50.86 | 15 mg olanzapine | 15 mg olanzapin |
| 46.33 | which has worsened | hvilket har forværret | 50.38 | the condition is quickly stabilized | tilstanden stabiliseres hurtigt |
| 45.67 | . 01 – | . 01 - | 49.70 | . recommended that | . anbefalet at |
| 45.65 | : known from | : kendt fra | 48.56 | course of treatment : quite | behandlingsforløb : ganske |
| 45.04 | has allegedly/reportedly been | har angiveligt været | 47.98 | which has worsened | hvilket har forværret |
| 44.80 | after 1 overnight stay | efter 1 overnatning | 47.90 | none/missing summary by | nihil resumé af |
| 44.29 | tells/reports that pt (patient) ., | fortæller pt ., | 47.13 | in the form of injectable | form af inj |
| 44.07 | discharged after | udskrives efter at | 47.02 | has allegdly/reportedly been | har angiveligt været |
| 43.87 | df141 ) (cocaine abuse) | df141 ) • | 46.71 | diagnosis note date • | diagnose noteringsdato • |
| 42.86 | 7 - 8 | 7 - 8 | 46.55 | : known from | : kendt fra |
| 42.44 | because he | grundet at han | 45.87 | doctor : none/missing | læge : nihil |
| 42.30 | 400 mg powder | 400 mg pulver | 45.81 | : none/missing summary | : nihil resumé |
| 41.96 | assesses/considers that the patient | vurderer at patienten | 45.68 | . 01 – | . 01 – |
| 41.57 | that the admission has | at indlæggelsen har | 45.39 | undifferentiated schizophrenia | udifferentieret skizofreni , |

*Top 25 **A**. unigrams, **B**. bigrams, and **C**. trigrams by **A**ttention **R**ollout score, from notes predicted as high risk of acute readmission (probability >=0.8), and including only the 10% most frequent n-grams globally.*

### Figure S14. Visualising label and prediction clusters with PCA

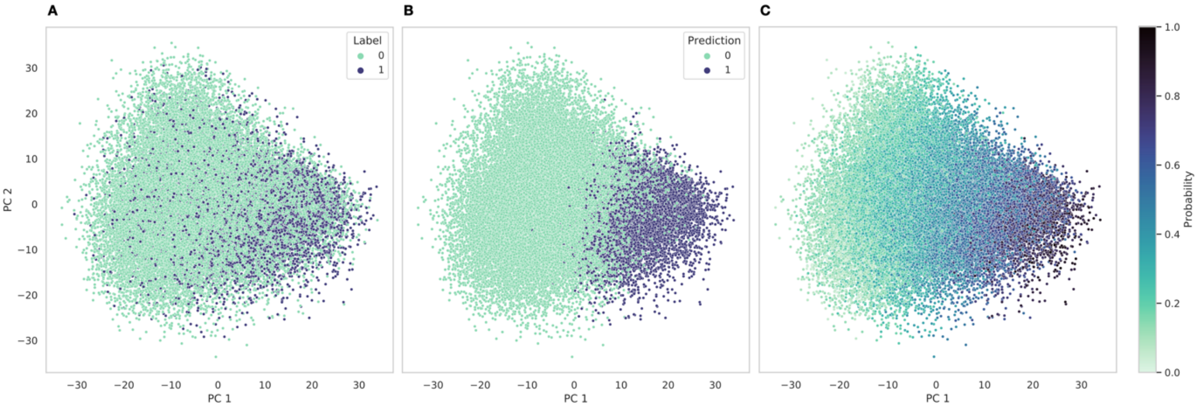


Principal Component Analysis with mean pooled embeddings (hidden states) of the discharge summary finetuning trainset. We took the mean of embedded notes within an admission before PCA and visualized each admission as one point and show here the relationship between the true labels and the model's (PsyRoBERTa) decision boundary. **A**. The label indicates whether the next admission was acute (1) or not acute (0). **B**. The prediction made for each admission when using 0.5 as threshold on the probability of the next admission being acute. **C**. The predicted probability of the next admission being acute.

### Figure S15. Acute readmission prediction performance of kNN classifiers

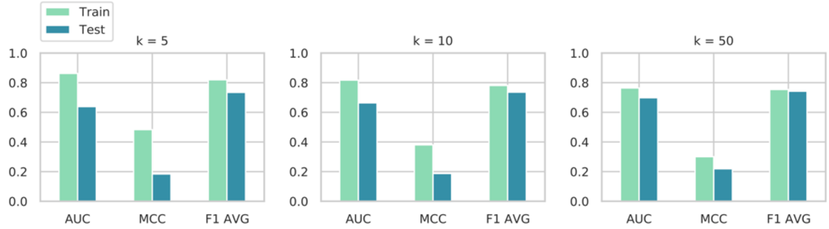


KNN classifiers were trained on PsyRoBERTa's representations of discharge summaries (visualized in **Figure S6**) to predict the acute readmission labels. The bar charts show train and test performance on three metrics (AUC, MCC and macro average $F_1$) and with K=5, 10 and 50.

### Figure S16. Visualising sex and age clusters with PCA

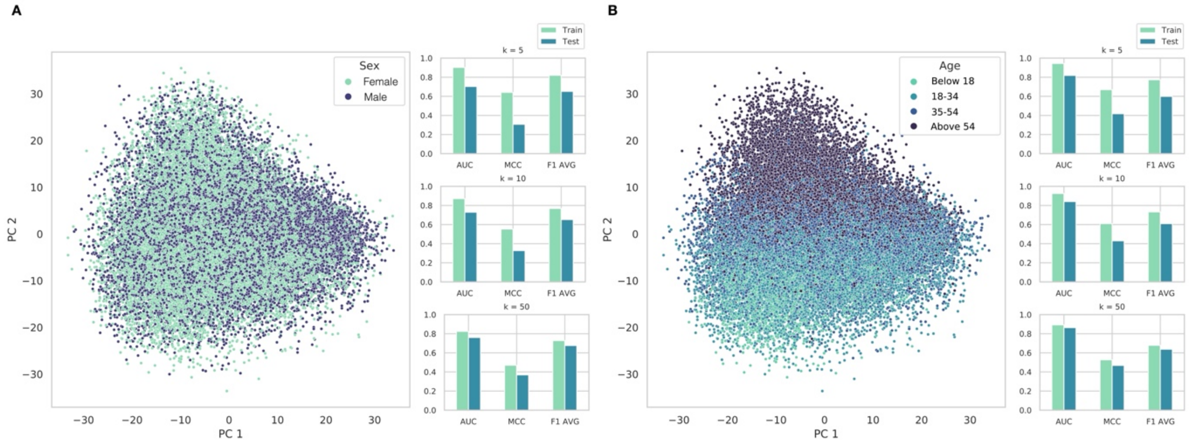


Principal Component Analysis with mean pooled embeddings (hidden states) of the discharge summary finetuning trainset. We took the mean of embedded notes within an admission before PCA and visualized each admission as one point. **A**. Each admission is colored by the sex of the patient, as it was recorded in the Electronic Health Records. The bar charts show train and test performance of three KNN classifiers (with K=5, 10 and 50) trained to predict sex. **B**. Each admission is colored by the age of the patient at the time of the admission. The bar charts show train and test performance of three KNN classifiers (K=5, 10 and 50) trained to predict age group.

#### Figure S17. Visualising clusters of the number of splits of discharge summaries (from data preprocessing) with PCA

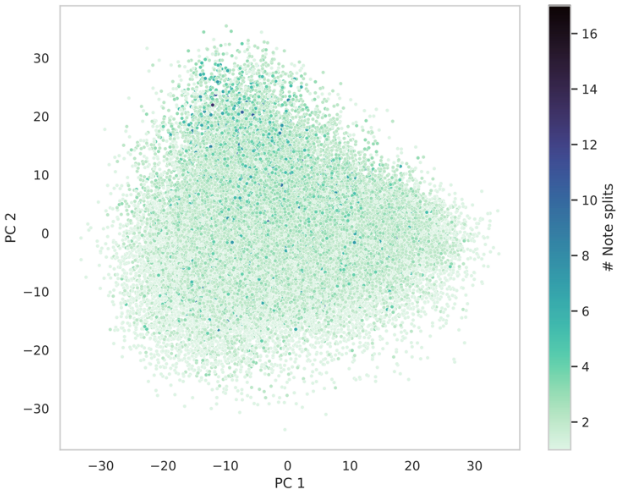


Principal Component Analysis with mean pooled embeddings (hidden states) of the discharge summary finetuning trainset. We took the mean of embedded notes within an admission before PCA and visualized each admission as one point. Each point is colored by the number of splits of the respective discharge summary. (Notes longer than 512 tokens were split into pieces of less than 513 tokens before finetuning.)

#### Figure S18. Diagnosis clusters visualized with PCA and diagnosis recognition performance of kNN classifiers

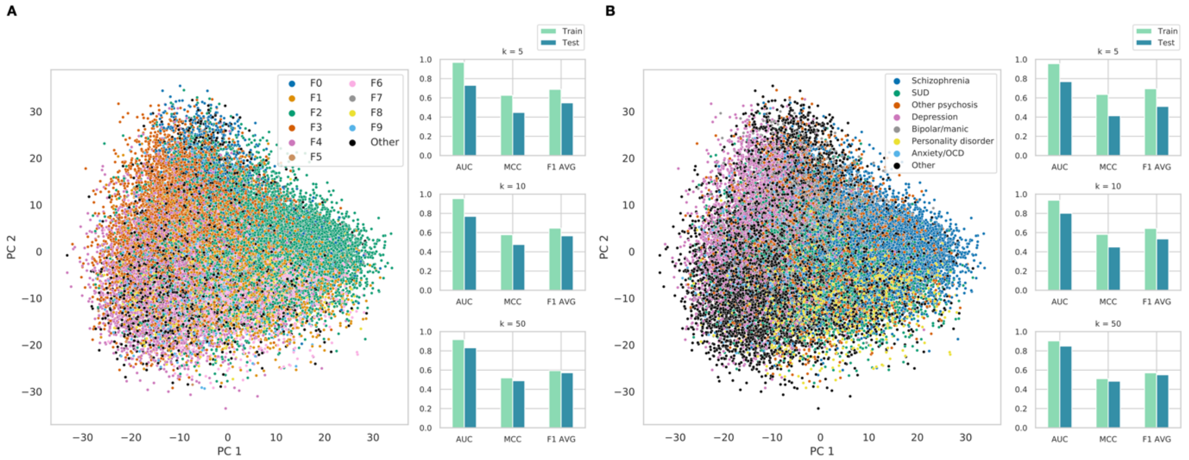


Principal Component Analysis with mean pooled embeddings (hidden states) of the discharge summary finetuning trainset. We took the mean of embedded notes within an admission before PCA and visualized each admission as one point. **A**. Each point is colored by psychiatric ICD codes (F0: Organic, including symptomatic, mental disorders; F1: Mental and behavioral disorders due to psychoactive substance use; F2: Schizophrenia, schizotypal and delusional disorders; F3: Mood [affective] disorders; F4: Neurotic, stress-related and somatoform disorders; F5: Behavioral syndromes associated with physiological disturbances and physical factors; F6: Disorders of adult personality and behavior; F7: Mental retardation; F8: Disorders of psychological development; F9: Behavioral and emotional disorders with onset usually occurring in childhood and adolescence). Although patients can have multiple diagnoses connected to the admission, we focused on the most severe diagnosis at the time of the admission.

#### Figure S19. Training and validation loss of language models finetuned for predicting acute readmission

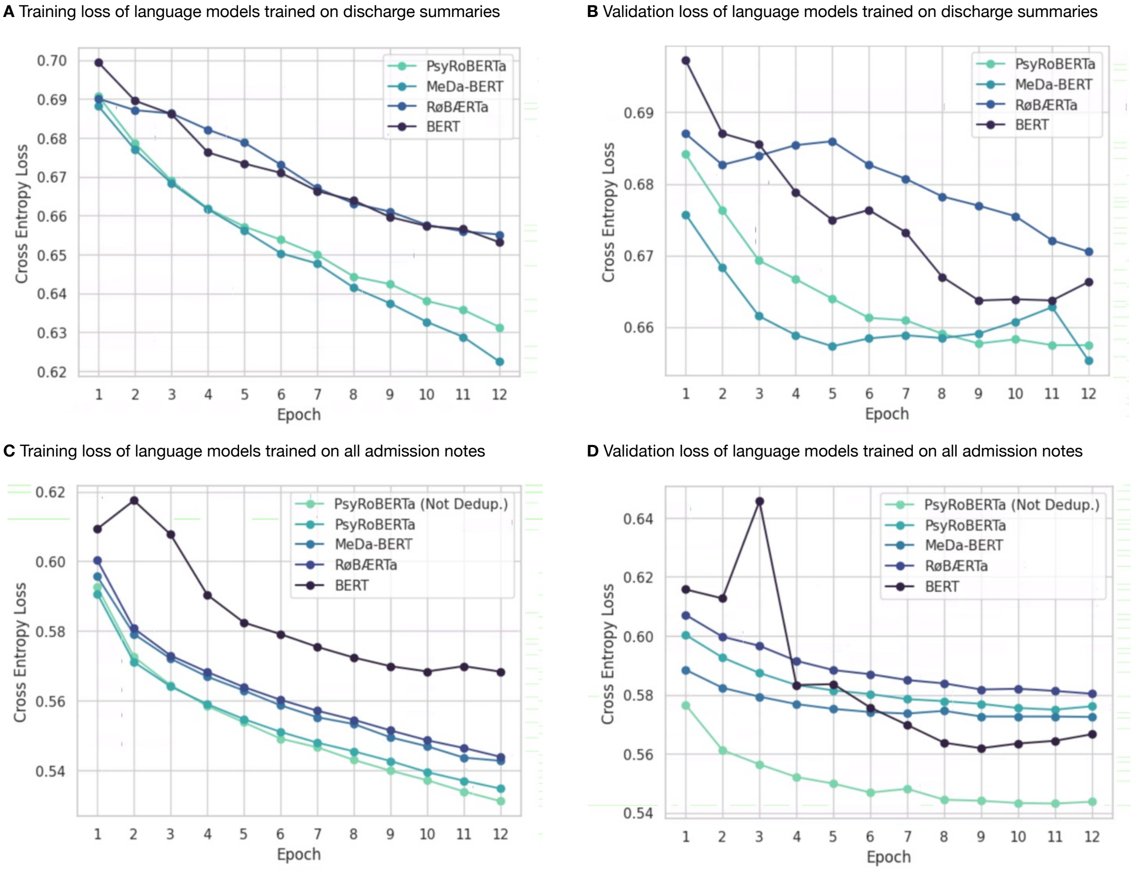


We show the mean loss across texts without aggregating on admissions. **A** and **B** show the training and validation loss of models finetuned on discharge summaries. **C** and **D** show the training and validation loss of models finetuned on all admission notes. For the latter, we finetuned on deduplicated admission notes (**eMethods**) but show the loss of PsyRoBERTa finetuned on the not deduplicated version of the data as well, for comparison. Although the validation loss is smaller when not deduplicating, the final predictive performance was greater when deduplicating (**Figure 3C**). This might be explained by the predictions being aggregated per admission, while the loss shown here is not.

#### Figure S20. Class clusters visualized with UMAP

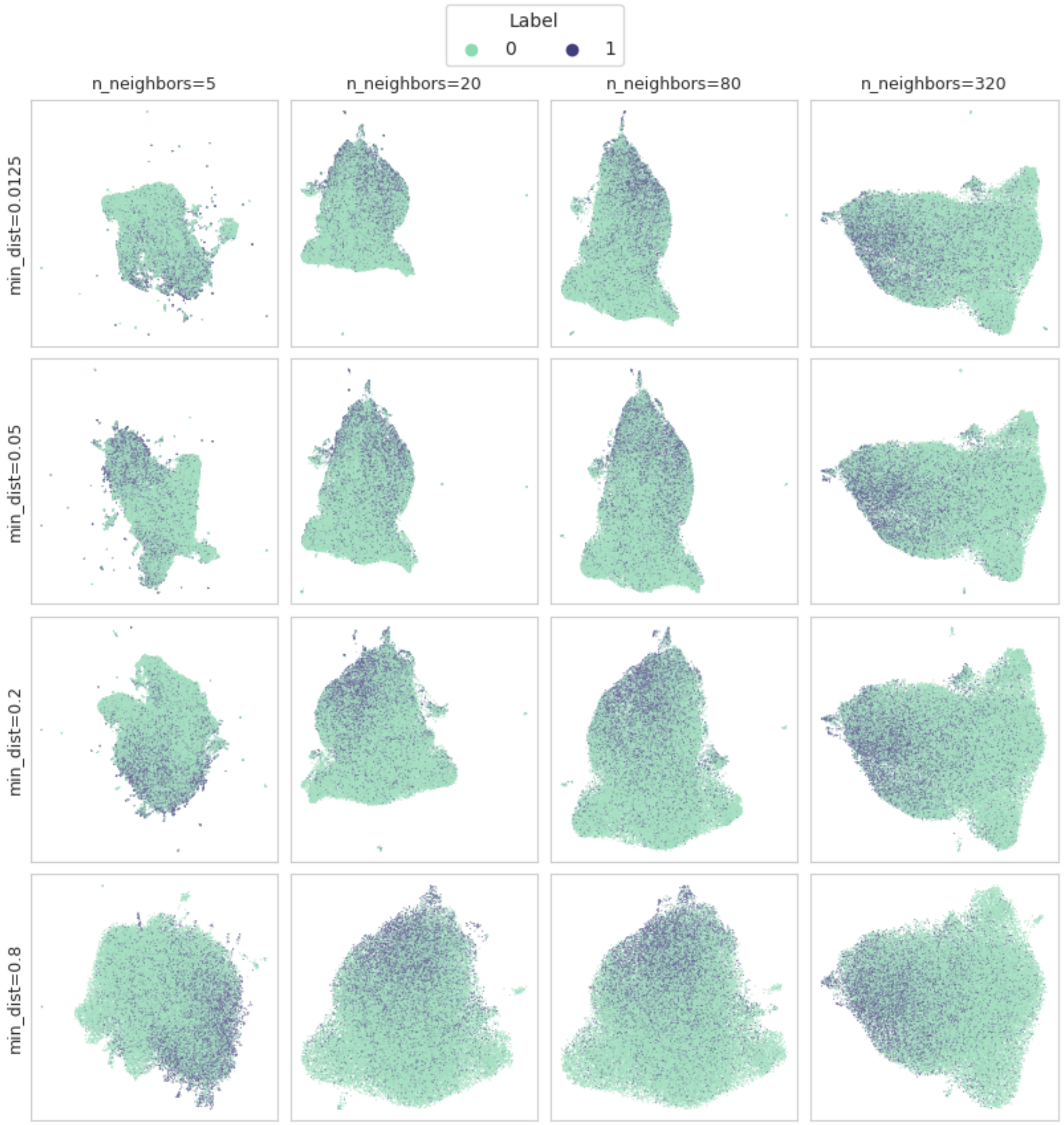


PsyRoBERTa representations of discharge summaries visualized with UMAP using cosine distance. Data points are colored by classes for the acute readmission prediction task.

#### Figure S21. Prediction clusters visualized with UMAP

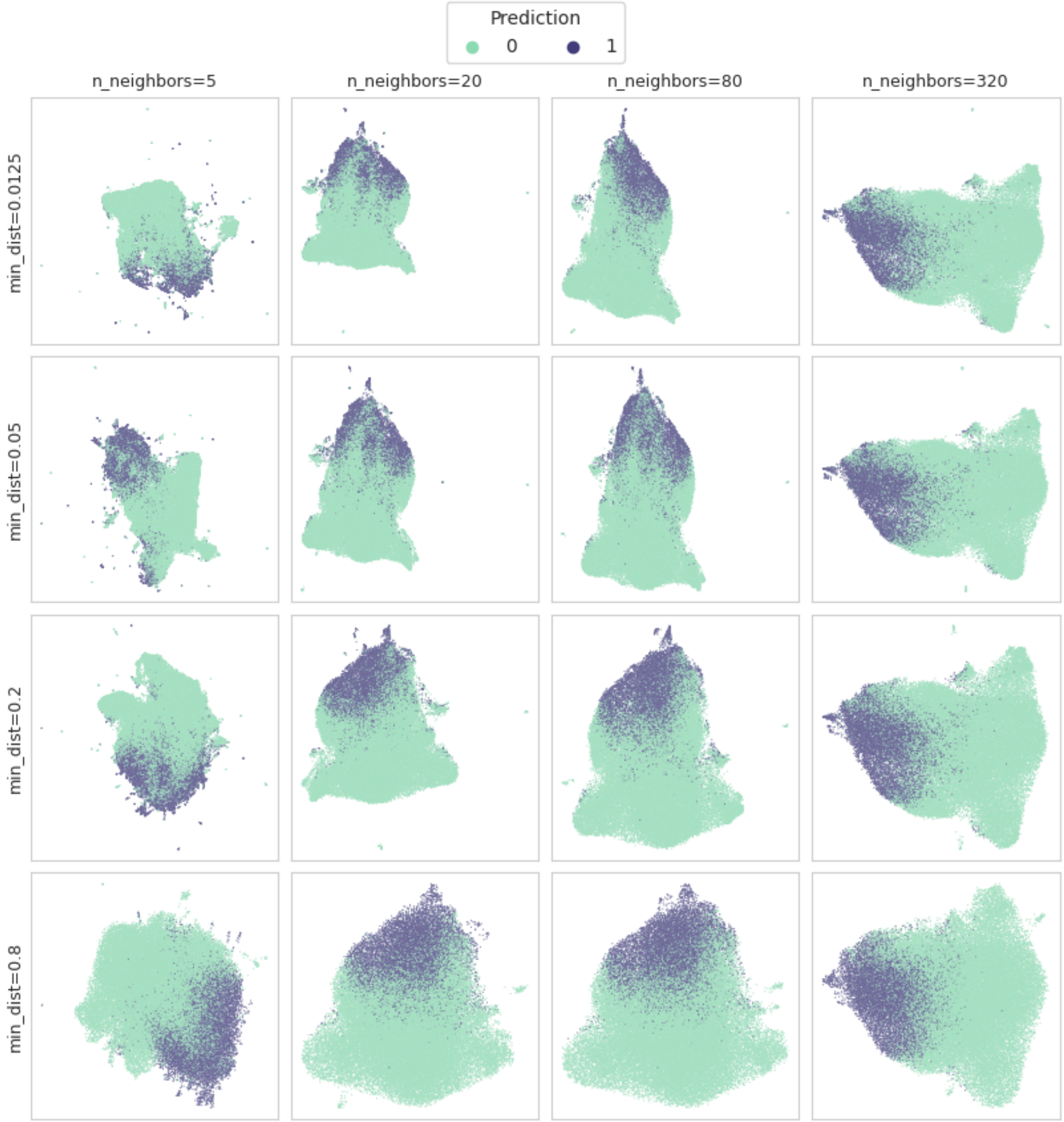


PsyRoBERTa representations of discharge summaries visualized with UMAP using cosine distance. Data points are colored by the prediction made for the acute readmission prediction task.

**Figure S22. Diagnosis clusters visualized with UMAP**

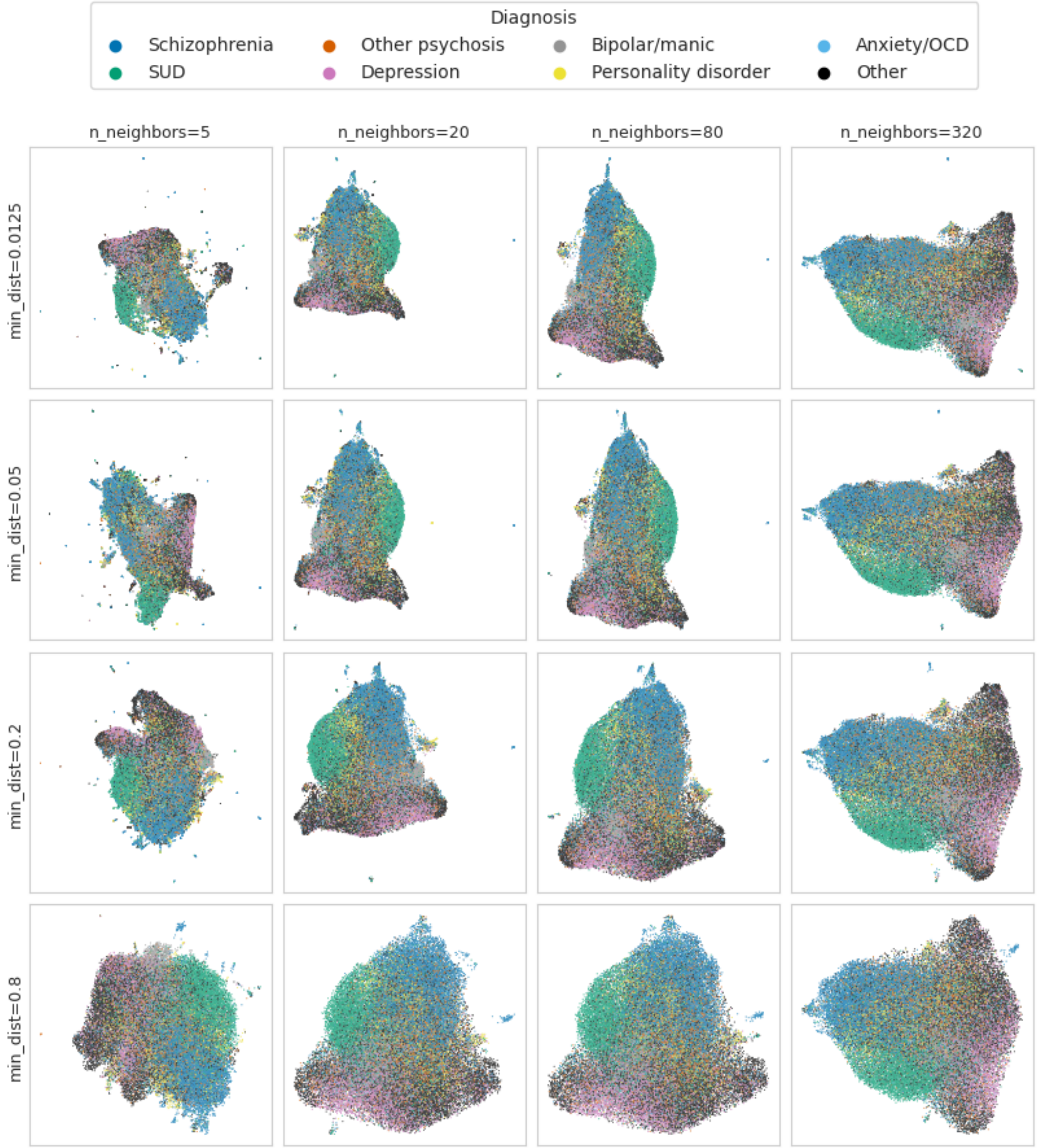


PsyRoBERTa representations of discharge summaries visualized with UMAP using cosine distance. Data points are colored by patients' main diagnosis at the time of the discharge.

#### Figure S23. ROC and Precision-Recall curves of language models

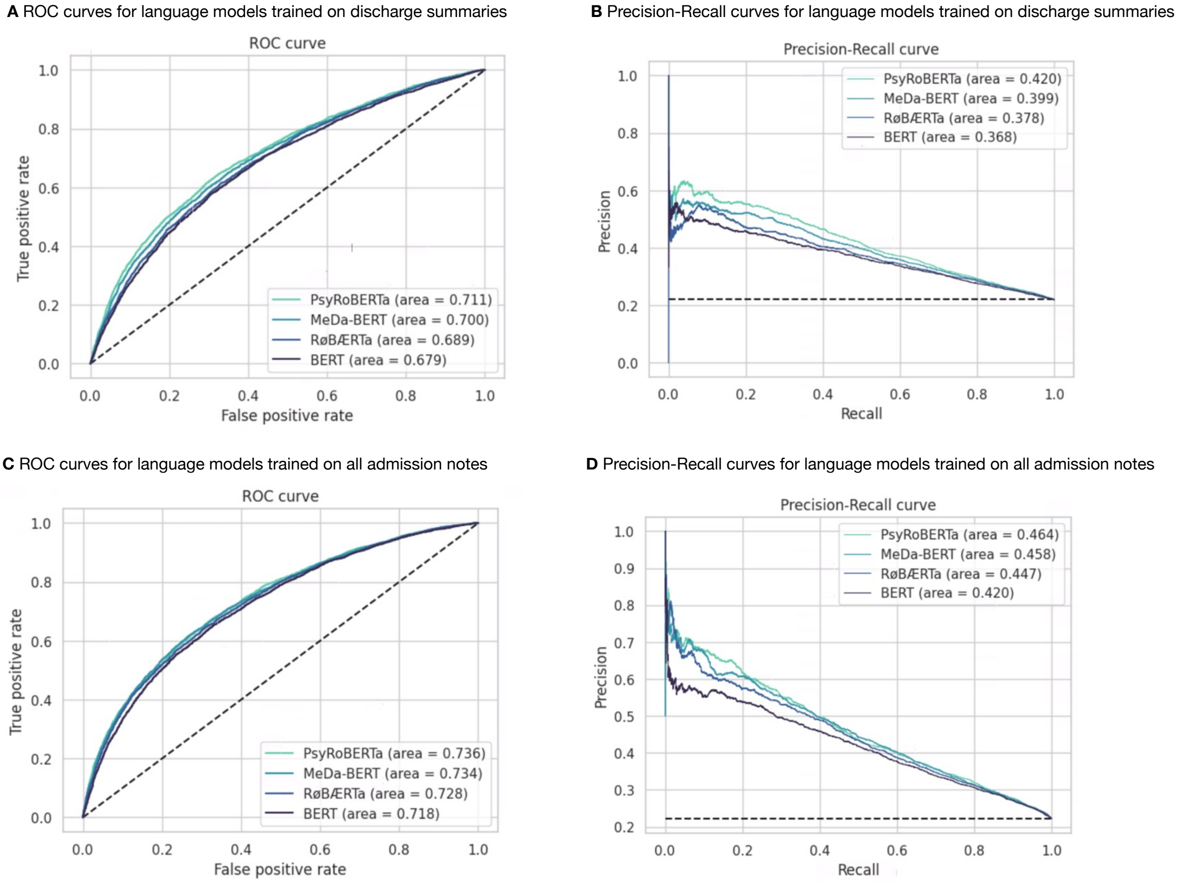


**A** and **B** show curves for models finetuned on discharge summaries, **C** and **D** show curves for models trained on all admission notes.

#### Figure S24. AUPRC and weighted $F_1$ of finetuned language models and logistic regression

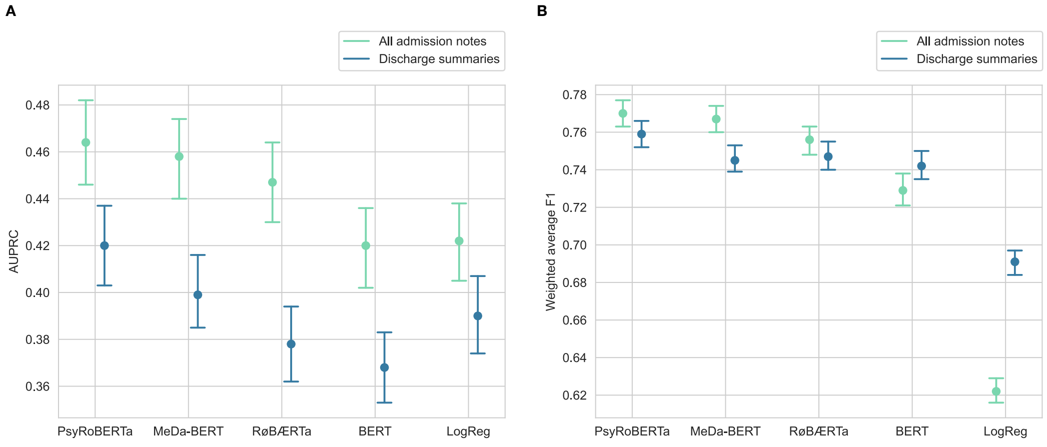


AURPRC (**A**) and weighted average $F_1$ scores (**B**) with 95% non-parametric bootstrap intervals.
